# Macrophage-augmented organoids recapitulate the complex pathophysiology of viral diseases and enable development of multitarget therapeutics

**DOI:** 10.1101/2025.02.27.25322840

**Authors:** Kuan Liu, Yining Wang, Jiajing Li, Jiahua Zhou, Ana Maria Gonçalves, Clara Suñer, Zhe Dai, Rick Schraauwen, Patrick P. C. Boor, Kimberley Ober-Vliegen, Francijna van den Hil, Dewy M. Offermans, Theano Tsikari, Ibrahim Ayada, Maikel P. Peppelenbosch, Martin E. van Royen, Monique M. A. Verstegen, Yijin Wang, Chloe M. Orkin, Harry L. A. Janssen, Valeria V. Orlova, Pengfei Li, Oriol Mitjà, Amaro Nunes Duarte-Neto, Luc J. W. van der Laan, Qiuwei Pan

## Abstract

The pathophysiology of viral diseases is complex, and often evokes strong inflammatory responses and tissue damage. Currently available *in vitro* models mainly recapitulate the viral life cycle *per se*, but fail to model immune cell-mediated pathogenesis. Here we build macrophage-augmented organoids (MaugOs) by integrating macrophages into organoids that are cultured from human liver tissues. We test the infections of two RNA viruses—hepatitis E virus (HEV) and SARS-CoV-2, and one DNA virus—monkeypox virus (MPXV), which either primarily or secondarily affect the human liver. In all three viral disease modalities, MaugOs recapitulate both infection and the resulting inflammatory response, *albert* to different levels. Intriguingly, this system showcases the ability to dissect the multifunctional role of human bile on HEV replication and inflammatory response through distinct mechanisms of action. MaugOs especially when integrated with pro-inflammatory macrophages recapitulate a prominent feature of inflammatory cell death triggered by HEV infection. Furthermore, we demonstrate a proof-of-concept in MaugOs to develop multitarget therapeutic strategies that simultaneously target the virus, inflammatory response, and the resultant inflammatory cell death.

## Introduction

The human population is continuously threatened by infectious diseases, in particular by a large number of epidemic and endemic pathogenic viruses. The liver has a unique immunological environment with essential functions in metabolism for example through the produced bile^1^. It serves as a preferential site for hepatotropic and non-hepatotropic virus invasions. Among the five classical hepatotropic viruses, hepatitis E virus (HEV) is the most common cause of acute viral hepatitis^2^. In addition, both clinical and experimental evidence has indicated susceptibility of the two major cell types in the liver, hepatocytes and cholangiocytes in particular, to SARS-CoV-2 infection^3–5^. A meta-analysis has reported a prevalence of liver injury of 40% in COVID-19 patients^6^. Although association of liver injury with monkeypox virus (MPXV) infection has been rarely investigated, a historical case report documented that virtually all cells in liver sections from a child who died from mpox were infected by the virus^7^.

Sustainable research on viral pathophysiology and development of novel treatment requires robust experimental models. Classically, immortalized cell lines have been used to model viral infections and test therapeutics^8^. More recently, 3D cultured organoids derived from adult stem cells or induced pluripotent stem cells have been increasingly explored as improved models for viral infections^9,10^. These *in vitro* cell line or organoid-based models are primarily restricted to recapitulating the life cycle of viruses *per se*, and thus fail to capture the complex pathophysiology of viral diseases in particular immune cell-mediated pathogenesis. Furthermore, therapeutics developed or tested in these cultures are exclusively antiviral regimens.

Severe viral diseases, such as COVID-19, mpox and viral hepatitis caused by SARS-CoV-2, MPXV and HEV respectively, have prominent features of pathological inflammation, leading to tissue damage in patients^11^. Macrophages have been recognized as a key, *albeit* not exclusive, type of immune cells orchestrating such hyperinflammation^12^. Macrophages residing in healthy tissues are either locally established before birth or recruited from circulating monocytes, whereas macrophages accumulating in infected tissues are mainly derived from circulating monocytes^12,13^. On the basis of function and activation status, macrophages can be classified into non-activated (M0), pro-inflammatory (M1), and anti-inflammatory (M2) subtypes^14^. Macrophages recognize invading pathogens to rapidly activate the inflammasome pathway^15^. Among the different types of human inflammasomes, the NLRP3 inflammasome is most relevant in recognizing viral infections^16^, which represents an emerging target for developing anti-inflammatory treatment^17^. For patients with severe viral infections, we reason that antiviral therapy alone is insufficient, but combining anti-inflammatory treatment is needed.

In this study, we established immune-augmented organoids by integrating macrophages into organoids derived from human liver tissues, termed as “macrophage-augmented organoids” (MaugOs). We employed two RNA viruses (HEV and SARS-CoV-2) and one DNA virus (MPXV) as disease modalities. We found that MaugOs support the productive infections of these three viruses, which in turn triggered inflammatory responses as well as inflammatory cell death. We further demonstrated that MaugOs can capture the multi-dimensions of viral pathophysiology, and enable the development of multitarget therapeutic strategies aiming at simultaneously targeting the virus and pathological inflammation, as well as the resultant inflammatory cell death.

## Results

### Establishing and characterizing MaugOs

To encompass infection and inflammation in one model, we integrated macrophages into organoids cultured from human liver tissues, namely intrahepatic cholangiocyte organoids (ICOs) originated from the intrahepatic biliary compartment^18^. We employed the human THP-1 monocyte cell line to differentiate into non-activated M0 macrophages (Extended Data Fig. 1a), generated primary macrophages from monocytes using peripheral blood mononuclear cells (PBMCs) isolated from healthy donors (Extended Data Fig. 1b-1d), and obtained macrophages differentiated from human induced pluripotent stem cells (hiPSCs)^19^. The hiPSC line expresses green fluorescent protein fused histone H2B (H2B-GFP), and this nuclear-localized GFP reporter is excellent for tracking and visualization (Extended Data Fig. 1e-1g). In addition, we have a unique access to human hepatic (tissue) macrophages isolated from liver graft preservation collected during liver transplantation at our institute (Extended Data Fig. 1h and 1i).

Organoids are cultured in Matrigel and thus immune cells are not capable of infiltrating into the hydrogel. To allow cell movement and integration, we performed a series of optimizations and technical innovations (details can be found in the methods section) to successfully establish MaugOs (Fig. 1a). Briefly, by lowering the concentration of Matrigel and seeding segmented ICOs with THP-1 macrophages, both cell types can gradually migrate and integrate into spheroids within 24 hours to form MaugOs (Fig. 1b and Supplementary Video S1). With this optimized protocol, MaugOs can be efficiently constructed using organoids derived from different donors (Fig. 1c and 1d). The MaugO morphology was further captured by optical imaging, immunofluorescent and H&E staining (Fig. 1e-1g, Supplementary Video 2 and 3). Genome-wide transcriptomic analysis revealed an intermediate pattern of MaugOs when compared with ICOs or macrophages alone based on the global gene expression profile (Fig. 1h).

**Fig. 1:**
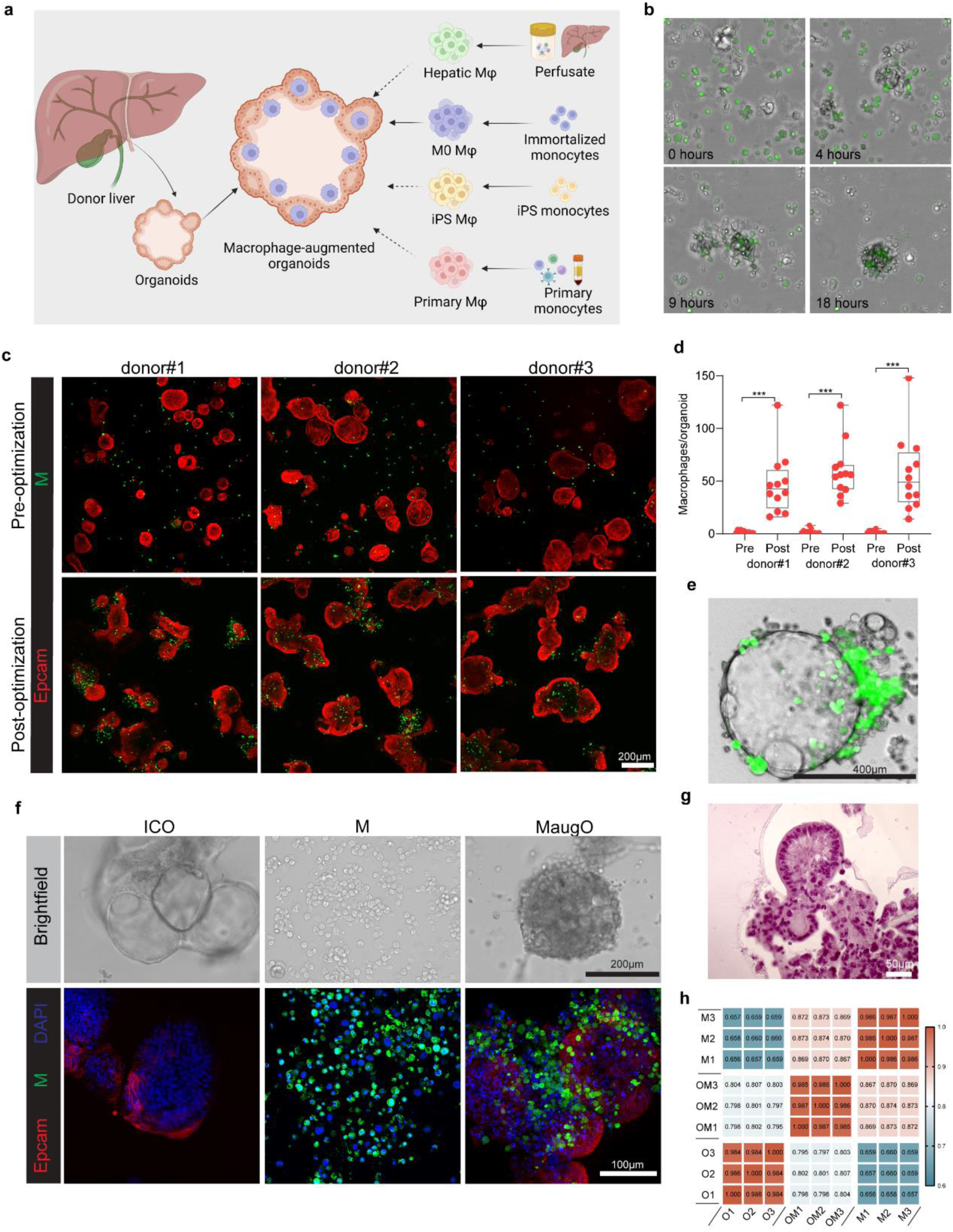
Establishment and morphological characterization of macrophage-augmented organoids. **a**, Schematic overview of constructing different types of macrophage-augmented organoids (MaugOs). The schematic illustration was generated in Biorender. **b**, Representative images of time–lapse confocal microscopy of MaugOs formation. Unstained organoid fragments (human ICOs) and CFSE pre-labelled macrophages differentiated from THP-1 monocytes were integrated for 24 hours. Images were taken every 0.5 hours (also see Supplementary Video S1). **c**, Immunofluorescence staining of MaugOs established from three different donor-derived organoids (donor #1, male; donor #2 and #3, female). Macrophages differentiated from THP-1 monocytes were labeled with CFSE (green), and organoids were stained with EpCAM (red). Images showing the comparison of MaugOs generated by the pre-optimized and post-optimized protocols. **d**. Quantification of the number of macrophages encapsulated in individual organoid. Statistical analysis comparing pre- and post-optimized protocols was performed by Mann–Whitney U test. n = 12; ***P <0.001. **e**, Brightfield images of MaguOs integrated with CFSE pre-labelled THP-1 macrophages for 2 days. **f**, Brightfield and immunofluorescence staining images depicting organoids (ICO),macrophages (M) and MaugOs. **g,** Hematoxylin and Eosin (H&E) staining of MaugOs.**h**, Correlation matrix of transcriptomic profiles of ICOs (O), macrophages (M), and MaugOs (OM) (n = 3). A value of 1 represents complete correlation, and a value of 0 represents no significant correlation.

Next, MaugOs were also established with primary monocyte-differentiated (Fig. 2a and 2b), hiPSC-derived (Fig. 2c and 2d) and hepatic (Fig. 2e) macrophages, demonstrating consistent efficiency of macrophage encapsulation in organoids (Extended Data Fig. 1j and 1k). The functionality of these established MaugOs was examined based on the response to bacterial lipopolysaccharide (LPS; a natural inflammatory stimulus) by quantifying a specified inflammatory gene panel including IL-1β, IL-6 and TNF-α. As expected, all types of tested macrophages but not organoids responded to LPS activation. Importantly, MaugOs can also effectively respond to LPS with the induction of most of the tested inflammatory genes, except for the MaugOs integrated with hepatic macrophages which only showed mild activation (Fig. 2f). We observed an unexpected phenomenon that direct contact of ICOs with hepatic macrophages, but not THP-1, primary monocyte-derived or hiPSC-derived macrophages, already triggered robust activation of inflammatory gene expression such as IL-1β, IL-6 and IL-12 (Extended Data Fig. 1l). Overall, optimizing technical approaches generated functional MaugOs that can respond to inflammatory stimulus.

**Fig. 2:**
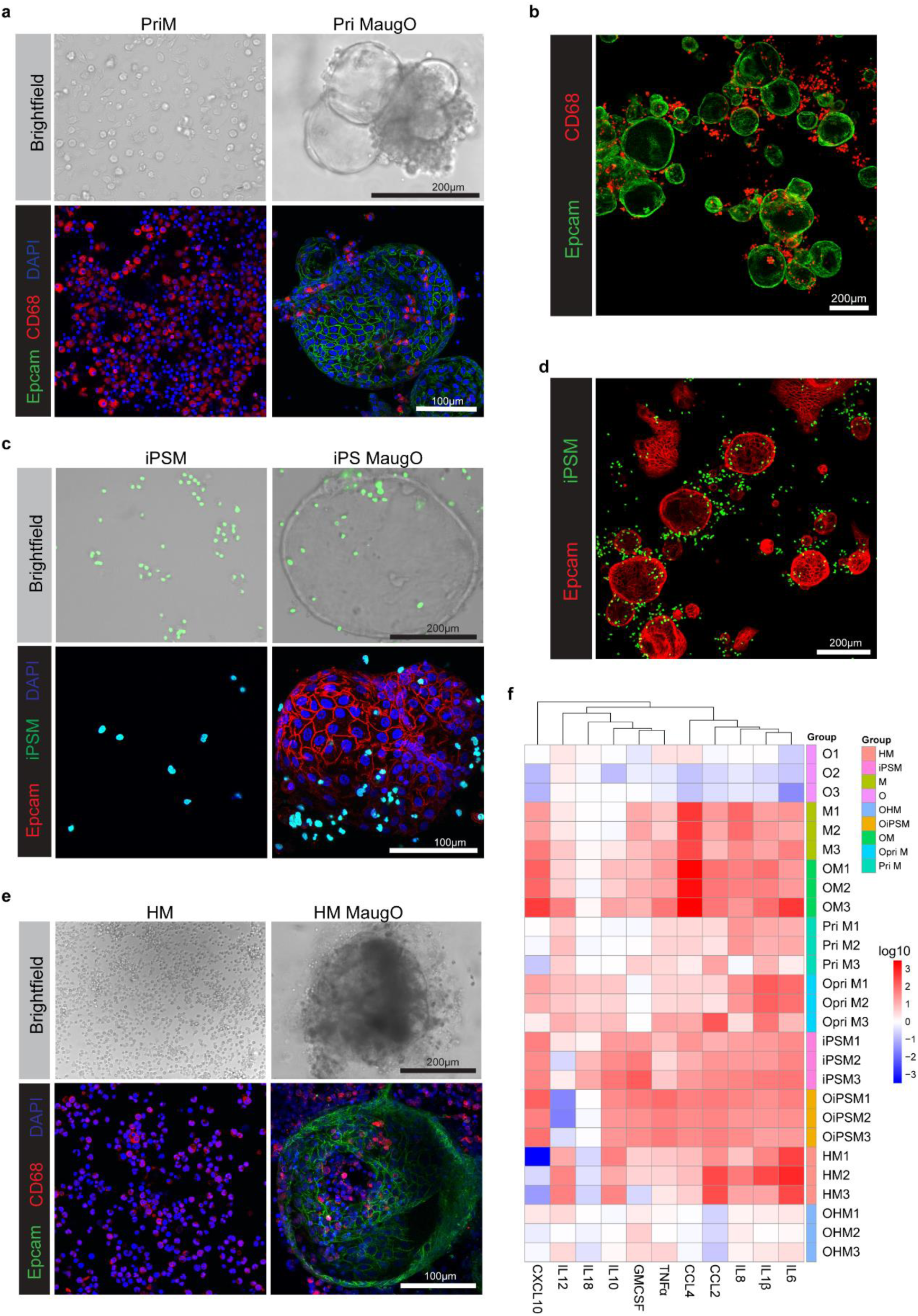
Morphological and functional characterization of macrophage-augmented organoids. **a**, Brightfield and confocal images of organoids integrated with primary macrophages differentiated from monocytes which were isolated from human PBMCs. Organoids were stained with EpCAM; macrophages were stained with CD68, a well-established macrophage marker. **b**, Immunofluorescence imaging overview of organoids integrated with primary macrophages. **c**, Brightfield and confocal images of hiPSC-derived macrophages (iPSM) and organoids incorporated with iPSM (iPS MaugO). **d**, Immunofluorescence imaging of iPS MaugO. **e**, Brightfield and immunofluorescence images illustrating hepatic macrophages (HM) and MaugOs integrated with hepatic macrophages (HM MaugO). **f**, The expression of inflammatory genes quantified by qRT-PCR upon stimulation of 1 μg/mL LPS for 12 hours. Each group was compared to the corresponding control without LPS treatment (n = 3). Primary macrophages (PriM) were generated from three independent buffy coats which are pools of human blood samples. Hepatic macrophages (HM1, HM2 and HM3) were derived from perfusates of three different donors. O1 - O3 (ICOs), OM1 - OM3 (MaugOs), Opri M1 - Opri M3 (Pri MaugOs), OiPSM1 - OiPSM3 (iPS MaugOs), OHM1 - OHM3 (HM MaugOs).

### MaugOs are susceptible to HEV and SARS-CoV-2 infections and trigger inflammatory responses

Both HEV and SARS-CoV-2 are RNA viruses. HEV is hepatotropic and can infect both hepatocytes and cholangiocytes in patient livers^20,21^. Histology analysis (Extended Data Fig. 2a) and immunofluorescent staining of HEV-infected patient liver sections with macrophage marker CD68 and the HEV ORF2 capsid protein showed their co-localization, suggesting HEV infection in liver tissue macrophages (Extended Data Fig. 2b-2c). In addition, we previously showed that ICOs support HEV infection^22^. The established MaugOs would therefore be suited to model HEV infection in culture. The common infection process is that a small amount of viruses first enter into the primary target cells, and then propagate, release and spread to other cell types such as immune cells^23^. To mimic this process, we first inoculated organoids with the virus, and then built MaugOs by further integrating macrophages (Fig. 3a). In MaugOs integrated with

**Fig. 3:**
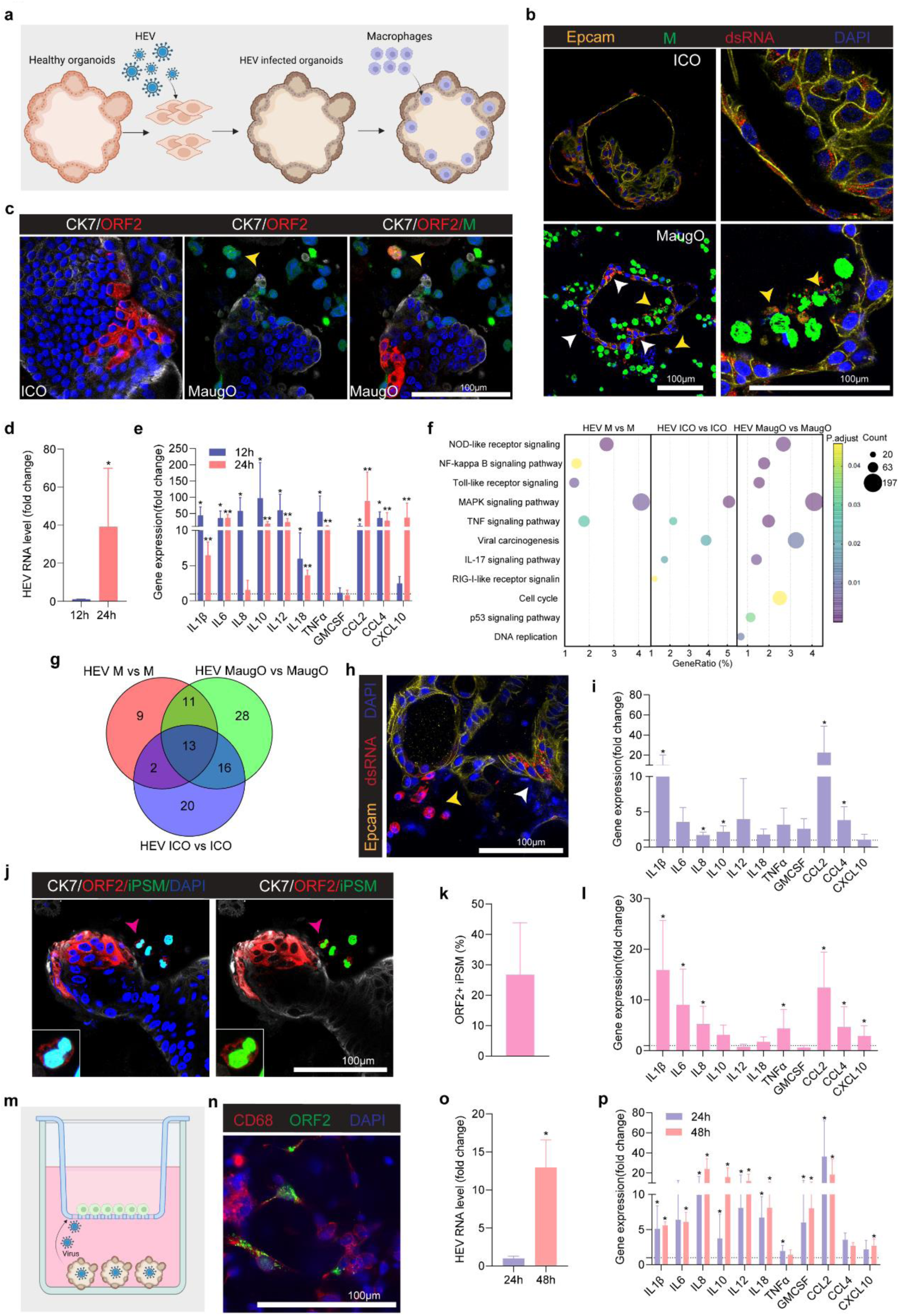
Macrophage-augmented organoids recapitulate HEV infection and inflammatory response. **a**, Schematic illustration of experimental design. **b,** Confocal imaging of viral double-stranded RNA (dsRNA) in HEV infected organoids (ICO) and infected organoids integrated with THP-1 macrophages (MaugO) for 48 hours. White arrows indicate infected organoids, whereas yellow arrows point to viral transmission to macrophages. **c**, Confocal images for HEV ORF2 protein (red) and epithelial marker CK7 (grey). Yellow arrows indicate the spread of HEV from organoids to macrophages in the infected MaugOs. **d** and **e,** Quantification of HEV viral RNA (n = 4) and inflammatory gene expression (n = 4-5) after HEV infected organoids integrated with THP-1 macrophages for 12 hours and 24 hours. The time point 12 hours served as control in panel **d** (normalized as 1). in panel **e**, the uninfected MaugOs served as control for the corresponding time points (normalized as 1). **f**, KEGG pathway analysis based on the genome-wide transcriptomic data showing the representative significantly regulated pathways for HEV infected macrophages (M), organoids (ICO), and MaugOs in comparison with their corresponding uninfected control groups (n = 3). Gene Ratio represents the proportion of genes from the dataset associated with a specific pathway in this analysis. **g**, Venn diagram depicting the number of significantly regulated pathways identified in KEGG analysis (n = 3). **h**, Immunofluorescence staining image of HEV infected organoids integrated with primary macrophages generated from monocytes isolated from PBMCs. White arrow shows HEV in organoids, and yellow arrow indicates viral transmission to macrophages. **i**, Quantification of inflammatory gene expression (n = 4) after HEV infected organoids integrated with primary macrophages for 24 hours. Uninfected MaugOs served as control (normalized as 1). **j** and **k**, Confocal imaging of HEV ORF2 protein (red) in MaugOs integrated with GFP-tagged hiPSC-derived macrophages, and quantification of ORF2 positive macrophages (n = 3). **l**, Quantification of inflammatory gene expression after HEV infected organoids integrated with hiPSC-derived macrophages for 24 hours (n = 4). Uninfected MaugOs served as control (normalized as 1). **m**, Schematic illustration of culturing HEV infected organoids with hepatic macrophages in a trans-well system. Organoids were cultured at the bottom, and hepatic macrophages were seeded in the 0.4 µm pore size insert. **n**, Immunofluorescence staining image showing ORF2 staining (green) in hepatic macrophages (CD68; red) after 48 hours culturing in the trans-well. **o** and **p**, Quantification of HEV viral RNA (n = 5) and inflammatory gene expression (n = 4) after HEV infected organoids cultured with uninfected hepatic macrophages for 24 hours and 48 hours. The time point 12 hour served as control in panel **o** (normalized as 1). In panel **p**, the uninfected groups served as control for the corresponding time points (normalized as 1). (**a** and **m**) The schematic illustration was generated in Biorender. (**d**, **e**, **i**, **l**, **o** and **p**) Data are mean ± SD. *p < 0.05; **p < 0.01; Mann-Whitney U test.

THP-1 monocyte-derived M0 macrophages, we observed HEV transmission from initially infected ICOs to macrophages, as shown by immunofluorescent staining of viral double-stranded RNA (dsRNA), the replicating intermediate (Fig. 3b and Extended Data Fig. 2d), and the ORF2 viral protein (Fig. 3c). Significant elevation by approximately 40-fold (n = 4; p < 0.05) of HEV RNA level was seen between 12 to 24 hours after inoculation of the virus to MaugOs, which indicates robust viral replication (Fig. 3d). Importantly, the expression of a panel of inflammatory genes (e.g. IL-1β, IL-6, IL-10, IL-12, TNF-α, CCL2 and CCL4) was dramatically increased at both 12 to 24 hours after MaugO assembly, compared to the uninfected MaugOs at the corresponding time points (Fig. 3e). This induction is recognized to be primarily driven by the integrated macrophages (Extended Data Fig. 3a) but hardly by the organoids (Extended Data Fig. 3b).

We performed genome-wide transcriptomic analysis in macrophages, organoids and MaugOs with and without HEV infection. Correlation analysis of the transcriptomes revealed an intermediate pattern of HEV infected MaugOs when compared with infected ICOs or infected macrophages, based on the global gene expression profile (Extended Data Fig. 2e). Pathway analysis revealed that the key pathogen-sensing and immune/inflammatory response related pathways are predominately regulated by HEV infection in macrophages and MaugOs but hardly in ICOs (Fig. 3f and Extended Data Fig. 2f-h). These include “NOD-like receptor signaling pathway”, “Toll-like receptor signaling pathway” and “NF-kappa B signaling pathway” (Fig. 3f). “Viral carcinogenesis” was significantly regulated in both ICOs and MaugOs but not in macrophages (Fig. 3f), suggesting a specific effect of HEV infection on organoid cells. “MAPK signaling pathway”, which is known to play a broad role in viral infection and inflammatory response^24^, was significantly regulated in all three groups (Fig. 3f). Furthermore, the number of significantly regulated pathways is substantially larger in MaugOs than in macrophages or ICOs (68 Vs. 35; 51), suggesting active interactions between macrophages and organoids upon HEV infection (Fig. 3g).

In some cases of hepatitis E, blood transfusion has been implicated as a source of HEV transmission^2^. Hypothetically, if viral titers are extremely high in contaminated blood products, these input viruses may simultaneously reach hepatic epithelial and immune cells. Thus, we also explored to first build MaugOs and then inoculate with HEV, to capture simultaneous exposure of organoids and macrophages to viral particles (Extended Data Fig. 3c). We first constructed ICO-based MaugOs, and comparatively profiled HEV infection along with the organoids and THP-1 macrophages (Extended Data Fig. 3d). HEV infection resulted in robust production of IL-1β and TNF-α cytokines in MaugOs but not in organoids (Extended Data Fig. 3e). We further validated using hepatocyte-like organoids differentiated from ICOs. Briefly, ICOs were cultured in previously defined hepatocyte differentiation medium, which subsequently acquired hepatic morphology of polygonal cell shapes and expression of hepatic markers such as albumin (Extended Data Fig. 3f and 3g)^22,25^. Hepatic organoids also support viral replication (Extended Data Fig. 3h), and ICO- and hepatic MaugOs are comparably susceptible to HEV infection (Extended Data Fig. 3i). HEV infection triggered significant production of IL-1β and TNF-α cytokines in hepatic MaugOs but not in hepatic organoids (Extended Data Fig. 3j). However, the levels appear to be substantially lower than those in ICO-based MaugOs (Extended Data Fig. 3e and 3j).

We next investigated HEV infection in ICO-based MaugOs integrated with primary macrophages differentiated from monocytes isolated from PBMCs using the original protocol (Fig. 3a). Similarly, we observed HEV transmission from infected organoids to macrophages based on staining of viral dsRNA (Fig. 3h). This triggered upregulation of inflammatory gene expression (e.g. IL-1β, CCL2 and CCL4) (Fig. 3i). We next evaluated hiPSC-derived macrophages, which are also susceptible to HEV infection (Extended Data Fig. 3k), resulting in induction of inflammatory gene expression (Extended Data Fig. 3l). In these MaugOs, transmission of HEV from infected ICOs resulted in over 25% of GFP and ORF2 double positive macrophages (Fig. 3j and 3k). This subsequently activated the expression of inflammatory genes including IL-1β, IL-6, IL-8, TNF-α and CCL2 (Fig. 3l). Finally, we also assessed HEV infection in hepatic macrophages isolated from liver organ perfusates, and found that these cells support HEV infection (Extended Data Fig. 3m) and trigger the expression of inflammatory genes, such as IL-1β, IL-6 and IL-8 (Extended Data Fig. 3n). Since hepatic macrophages obtained from graft preservation fluid are already activated during integration with organoids (Extended Data Fig. 1l), we found that HEV inoculation largely failed to further activate inflammatory response in MaugOs integrated with hepatic macrophages (Extended Data Fig. 3o), which is consistent with observations when exposed to LPS (Fig. 2f). We therefore explored an alternative approach by co-culturing ICOs with hepatic macrophages in a trans-well system (Fig. 3m). Interestingly, HEV transmission from organoids to macrophages occurred through the paracrine route (Fig. 3n), and there was robust viral replication showing significantly elevated HEV RNA level by approximately 10-fold (n = 5; p < 0.05) when comparing the 24 versus 48 hour time point (Fig. 3o). The expression of a large number of inflammatory genes (e.g. IL-1β, IL-8, IL-10, IL-12 and CCL2) was significantly upregulated at both 24 and 48 hour time points (Fig. 3p).

Although primarily targeting the lung, SARS-CoV-2 can also infect the liver, especially cholangiocytes due to high expression of the receptor ACE2^4,26^. In our established MaugOs challenged with SARS-CoV-2, we observed viral transmission from infected ICOs to macrophages shown by staining of viral dsRNA (Extended Data Fig. 4a) and viral protein (Extended Data Fig. 4b). The infection in MaugOs was also confirmed by qRT-PCR quantification of viral RNA (Extended Data Fig. 4c). This resulted in moderate activation of inflammatory gene expression (Extended Data Fig. 4d), and significantly elevated production of IL-1β (n = 6; p < 0.01) (Extended Data Fig. 4e). Collectively, MaugOs are capable of recapitulating the infections of HEV and SARS-CoV-2, and the resulting inflammatory responses.

### MaugOs incorporated with polarized macrophages recapitulate HEV infection, inflammatory response and inflammatory cell death

Non-activated M0 macrophages can be polarized into M1 pro-inflammatory or M2 anti-inflammatory subtypes^14^. We generated these three subtypes from THP-1 monocytes using well-established protocols^27^, and confirmed their phenotypes by morphological visualization and cell surface marker analysis (Extended Data Fig. 5a-5c). We constructed MaugOs using these macrophage subtypes to assess their responses to HEV infection (Fig. 4a). Quantification of inflammatory gene expression (e.g. IL-1β, IL-6, TNF-α, CCL2 and CCL4) revealed that M1-intergrated MaugOs are the most sensitive to HEV infection showing the highest levels of induction of these genes (Fig. 4b). This is also reflected by the highest level of IL-1β secretion (approximately 800 Vs. 500 pg/mL) (Fig. 4c).

**Fig. 4:**
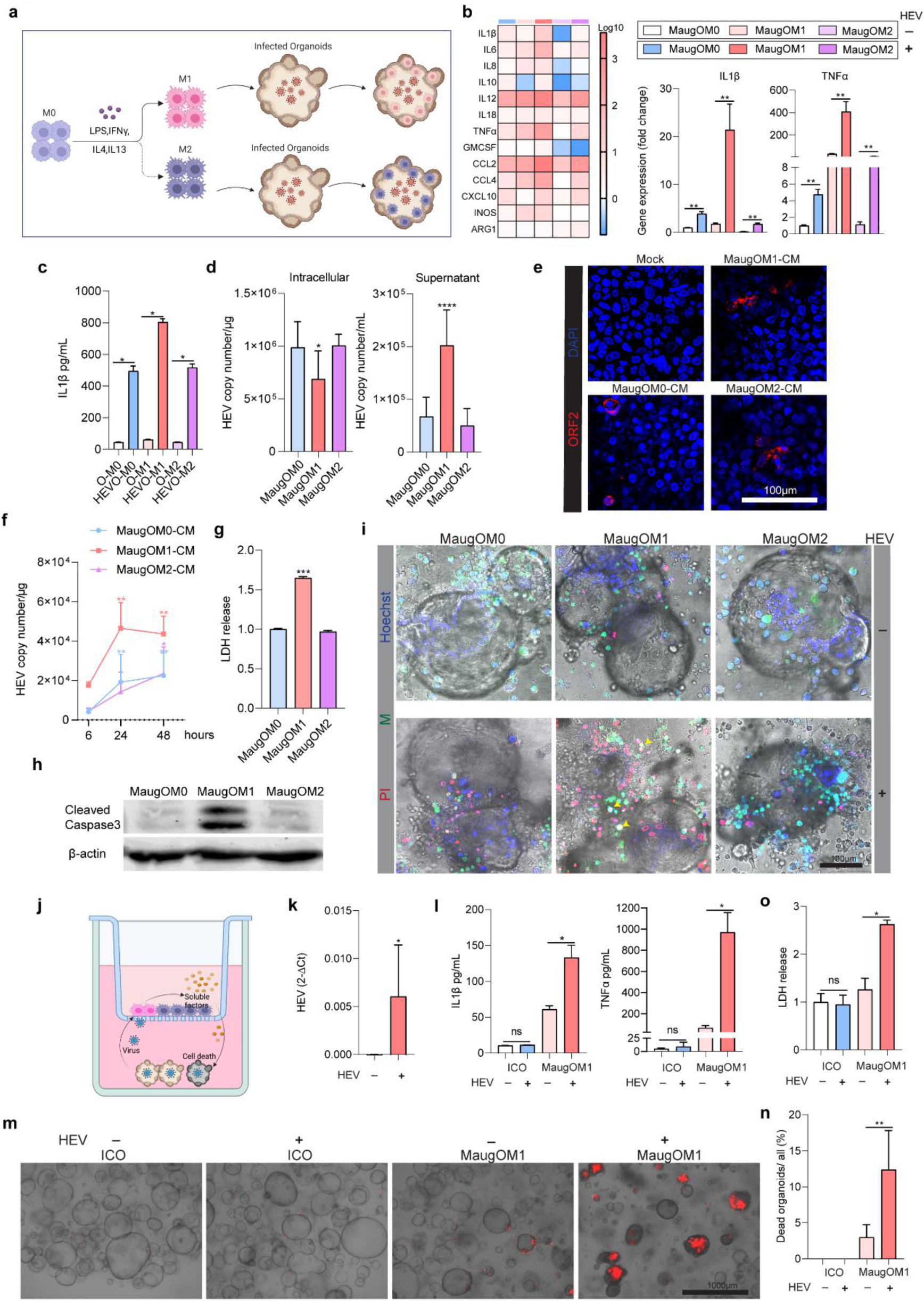
Organoids augmented with polarized macrophages recapitulate HEV infection, inflammatory response and inflammatory cell death. **a**, Schematic illustration of the differentiation processes of M1/M2 macrophages from THP-1 monocytes and the integration with HEV infected organoids. **b**, Quantification of inflammatory gene expression in uninfected organoids integrated with M0/M1/M2 macrophages and HEV infected organoids integrated with M0/M1/M2 macrophages (MaugOM0/MaugOM1/MaugOM2) for one day. The uninfected organoids integrated with M0 macrophages served as control (normalized as 1). Heatmap shows the average of relative gene expression in samples (n = 2-5). IL-β and TNF-α were shown individually (n = 5). **c**, Quantification of IL-β cytokine production by ELISA (n = 4). **d**, Quantification of intracellular and supernatant HEV RNA levels by qRT-PCR (n = 12). **e**, Immunofluorescence staining images of HEV ORF2 protein expression (red) in human liver Huh7 cells inoculated with the conditional medium (CM) harvested from HEV infected MaugOs integrated with M0, M1 or M2 macrophages (MaugOM0-CM, MaugOM1-CM, MaugOM2-CM). **f**, Quantification of HEV viral RNA in Huh7 cells inoculated with the corresponding supernatant (n = 6). **g**, Quantification of lactate dehydrogenase (LDH) release in HEV infected MaugOs integrated with M0, M1 or M2 macrophages (n = 8). MaugOM0 served as control (normalized as 1). **h**, Western blotting analysis detecting the cleaved caspase 3 in three types of HEV infected MaugOs. **i**, Confocal microscopy imaging of Propidium Iodide (PI; red) staining, and CFSE pre-labelled M0/M1/M2 macrophages (green). Arrows point to representative macrophages with PI and CFSE double positivity. **j**, Schematic illustration of culturing HEV infected organoids with M1 macrophages in a trans-well system. Organoids were cultured at the bottom, and M1 macrophages were seeded in the 0.4 µm semipermeable insert. **k**, qRT-PCR quantification of HEV RNA level in M1 macrophages (n = 4). Samples were cultured for 2 days and harvested from the insert. **l**. Quantification of IL-β and TNF-α cytokine production in supernatant by ELISA (n = 4). Organoids (ICO) and organoids co-cultured with M1 macrophages (MaugOM1) infected with or without HEV. **m** and **n**, Visualization and quantification of cell death in organoids using Propidium Iodide (PI; red) staining marking dead or dying cells (n = 4-5). **o**, Measurement of lactate dehydrogenase (LDH) levels (n = 4). ICO without HEV infection served as control (normalized as 1). (**b**, **c**, **d**, **f**, **g**, **k**, **l**, **o** and **n**) Data are mean ± SD. *p < 0.05; **p < 0.01; Mann-Whitney U test. (**a** and **j**) Schematic illustration was generated by biorender.

Quantification of HEV RNA revealed striking results that MaugOs integrated with the pro-inflammatory M1 macrophages had the lowest level of intracellular viral RNA level, while produced the highest level in the supernatant (Fig. 4d). We confirmed the infectiousness of produced HEV virus particles by inoculating the human liver cell line Huh7, as shown by the expression of ORF2 viral protein (Fig. 4e). We further quantitatively determined the increased viral RNA levels in Huh7 cells upon inoculation of the corresponding supernatant (Fig. 4f), which again confirmed the highest level of virus release from the M1 macrophage integrated MaugOs. We postulate that the pro-inflammatory M1 macrophages have a better capacity in triggering inflammatory cell death upon HEV infection, leading to increased release of the virus into supernatant. To confirm this, we measured lactate dehydrogenase (LDH) release, a classical indicator of cellular damage^28^. Indeed, HEV infection triggered the highest level of LDH release in M1 macrophage integrated MaugOs (about 1.5-fold compared to that in M0 or M2 based MaugOs) (Fig. 4g). This was further confirmed by Western blotting of cleaved caspase-3 (Fig. 4h), a marker for detecting apoptotic cells. Staining based on positivity of the propidium iodide (PI) dye showed that HEV infection triggered cell death predominantly occurred in organoids, but also in macrophages to some extent, and consistently such inflammatory cell death is most prominent in the M1 subtype MaugOs (Fig. 4i).

To further characterize, we co-cultured the pro-inflammatory M1 macrophages with HEV-infected organoids in a trans-well system (Fig. 4j). This resulted in viral transmission to macrophages (Fig. 4k), and inflammatory response as indicated by significantly elevated production of IL-1β and TNF-α cytokines (Fig. 4l). Intriguingly, the percentage of dying organoid cells (Fig. 4m and 4n) and the resultant LDH release (Fig. 4o) were dramatically increased. Of note, HEV infection *per se* has minimal effect on cell death in organoids, if without the presence of macrophages (Fig. 4m and 4n). These results demonstrate that macrophages mediate HEV-triggered inflammatory cell death, which is at least partially through a paracrine mechanism. Collectively, in addition to viral replication and inflammatory response, our model can also capture immune cell mediated tissue damage.

### Capturing the multifunctional role of human bile on HEV replication and inflammatory response in MaugOs

Bile is a potent digestive surfactant facilitating absorption of nutrients and is produced by hepatocytes in the liver. Bile mainly consists of bile acids, which also have hormonal actions mainly through the farnesoid X receptor (FXR) and Takeda G protein-coupled receptor 5 (TGR5) (Fig. 5a)^29^. HEV is primarily transmitted through a fecal-oral route. Because it is excreted from the infected liver via the biliary tract into the feces, HEV is exposed to high concentrations of bile acids prior to release into the intestine^30^. In ICOs harboring the HEV subgenomic replicon coupled to a luciferase reporter (Extended Data Fig. 6a), treatment with human bile dose-dependently inhibited viral replication-related luciferase activity (Fig. 5b). This was further confirmed in ICOs harboring the fully infectious HEV genome (Fig. 5c). We next tested the two most common types of primary bile acids, cholic acid (CA) and chenodeoxycholic acid (CDCA)^31^. Consistently, both CDCA and CA significantly inhibited HEV replication in the replicon (Extended Data Fig. 6b) and in the infectious model (Fig. 5d). Of note, the concentrations of human bile as well as the representative bile acids that are used in this study had minimal effects on the growth of organoids based on brightfield imaging and cell viability assay (Extended Data Fig. 6c and 6d). This anti-HEV effect was completely reversed in both HEV models, when co-treated with z-guggulsterone (ZGG)—an FXR antagonist (Fig. 5e), suggesting that the antiviral mechanism is primarily through the FXR pathway. In HEV infected THP-1 macrophages, exposure to human bile also inhibited HEV replication and inflammatory gene expression (e.g. IL-1β, IL-8 and TNF-α) (Extended Data Fig. 6e-6h).

**Fig. 5:**
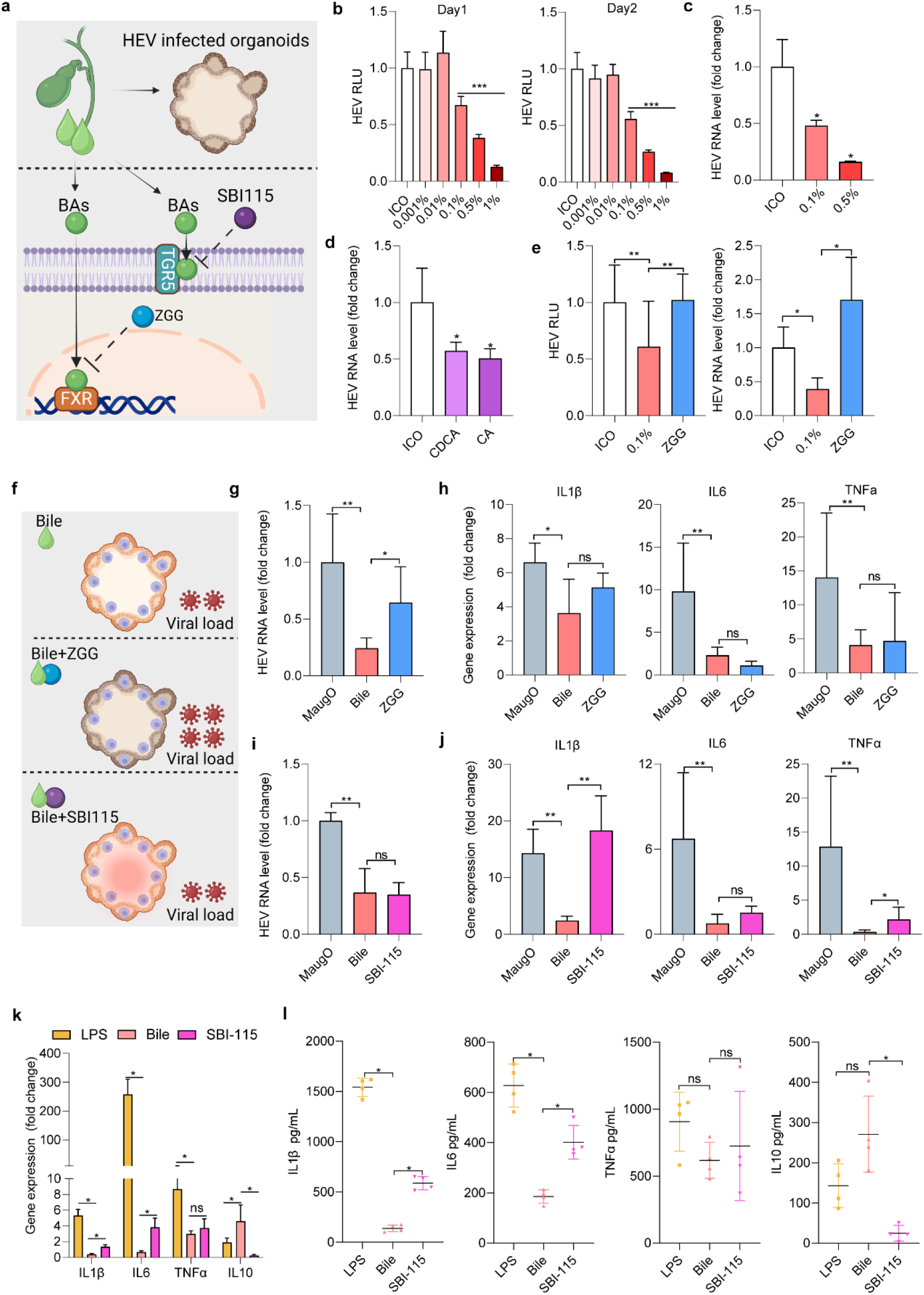
Human bile on HEV infection and inflammatory response. **a**, Schematic illustration of the experimental design (upper panel) and bile acids signaling pathway with inhibitors targeting the key receptors (lower panel). **b**, The effects of human bile treatment with a series of concentrations for one or two days in organoids (ICO) harboring the HEV subgenomic replicon (n = 8). RLU, related luciferase units. **c**, Bile treatment for two days on HEV RNA level quantified by qRT-PCR in organoids (ICO) harboring infectious HEV (n = 4). **d**, Treatment with bile acids (100 μM CDCA or 100 μM CA) for two days on HEV RNA level in organoids (ICO) with the HEV infectious model (n = 4) **e**, The effects of 10 μM z-guggulsterone (ZGG) co-treated with 0.1% bile for two days on viral replication related luciferase activity in the subgenomic replicon (left, n = 10) or HEV RNA level in the infectious model (right, n = 4). **f**, Schematic overview of bile co-treated with ZGG or SBI-115 in MaugOs on HEV infection and inflammatory response. **g** and **h**, Quantification of HEV viral RNA (n = 6) and inflammatory gene expression (n = 6) in HEV infected MaugOs treated with 0.1% bile or co-treated with 10 μM ZGG. **i** and **j**, Quantification of HEV viral RNA (n = 6) and inflammatory gene expression in HEV infected MaugOs treated with 0.1% bile or co-treated with 50 μM SBI-115 (n = 6) **k** and **l**, Quantification of gene expression (n = 4) and cytokine production (n = 4) of IL-β, IL-6, TNF-α and IL-10 in 1 μg/mL LPS incubated MaugOs treated with bile or co-treated with SBI-115 for one day. Data are presented as mean ± SD. *p < 0.05; **p < 0.01; Mann-Whitney U test. (**a** and **f**) The schematic illustration was generated in Biorender.

To simultaneously assess the role of bile on viral infection and the inflammatory response, we next employed MaugOs integrated with THP-1 macrophages (Fig. 5f). Similarly, human bile inhibited HEV replication in MaugOs, and this effect was abrogated by ZGG (Fig. 5g). In parallel, we found that bile can also inhibit HEV-triggered inflammatory gene expression, but this effect was not affected by ZGG treatment (Fig. 5h). To specifically probe the anti-inflammatory mechanism of bile, we treated HEV-infected MaugOs with SBI-115—a TGR5 antagonist. Interestingly, SBI-115 had no effect on the anti-HEV activity of bile shown by quantification of viral RNA (Fig. 5i). However, the inhibitory effects of bile on the expression of inflammatory genes (e.g. IL-1β and TNF-α) was completely or partially reversed by SBI-115 treatment (Fig. 5j). To validate the specificity as well as broaden the implications in inflammatory response, we further investigated in MaugOs activated by LPS. Similarly, human bile inhibited the expression and production of pro-inflammatory effectors, and this was abrogated by SBI-115 treatment (Fig. 5k and 5l). In parallel, bile elevated the level of an anti-inflammatory effector (IL-10), which was abrogated by SBI-115 treatment (Fig. 5k and 5l). These effects on inflammatory response were not affected by ZGG treatment (Extended Data Fig. 6i and 6j). Collectively, these results demonstrated that MaugOs are capable of capturing the anti-HEV and anti-inflammatory effects of human bile (partially) through FXR and TGR5 pathways, respectively.

### Therapeutically targeting the NLRP3 inflammasome and combination of antiviral with anti-inflammatory treatment

We have previously shown that HEV strongly activates the NLRP3 inflammasome in macrophages, representing a key mechanism of driving pathological inflammation^32^. Using the three indicated pharmacological inhibitors targeting the NLRP3 inflammasome cascade (Fig. 6a), we demonstrated that these inhibitors had minimal effects on HEV replication in MaugOs (Fig. 6b) but affected the expression of inflammatory genes (Extended Data Fig. 5d). Importantly, they completely abolished HEV-triggered IL-1β secretion (the hallmark of inflammasome activation) in MaugOs integrated with M0 macrophages (Fig. 6c). This is consistent with Western blotting results on protein levels of secreted mature IL-1β in supernatant (Fig. 6d). Treatment with these pharmacological inhibitors also profoundly inhibited HEV-triggered IL-1β secretion in MaugOs integrated with the pro-inflammatory M1 macrophages (Fig. 6e). Because of the occurrence of hyperinflammation in severely infected patients, we reason that multitarget therapy by combining antiviral and anti-inflammatory treatment is necessary to achieve a favorable clinical outcome^15,33^. To demonstrate a proof-of-concept, we first tested the combination of ribavirin (a clinically used antiviral drug for treating hepatitis E) and MCC950 (an NLRP3 inhibitor) in HEV-infected MaugOs integrated with M0 macrophages (Fig. 6f). The combination resulted in simultaneous inhibition of viral replication and IL-1β secretion, whereas ribavirin or MCC950 alone only inhibited HEV replication or IL-1β production respectively (Fig. 6G and 6h). No major effects were observed on TNF-α (Fig. 6i) and IL-10 production (Extended Data Fig. 5e). We then tested the combination of ribavirin with dexamethasone, a clinically used anti-inflammation drug which is known to inhibit virus-triggered NLRP3 inflammasome^15,34^. Consistently, this combination significantly inhibited HEV replication (Fig. 6j), and IL-1β gene expression (Extended Data Fig. 5f) and production (Fig. 6k). Similar results were observed on TNF-α expression (Extended Data Fig. 5g) and secretion (Fig. 6l). Interestingly, the level of an anti-inflammatory cytokine IL-10 was significantly elevated by the combination treatment (Extended Data Fig. 5h). Ribavirin alone inhibited viral replication only, while dexamethasone alone regulated inflammatory effectors only (Fig. 6g-6l).

**Fig. 6:**
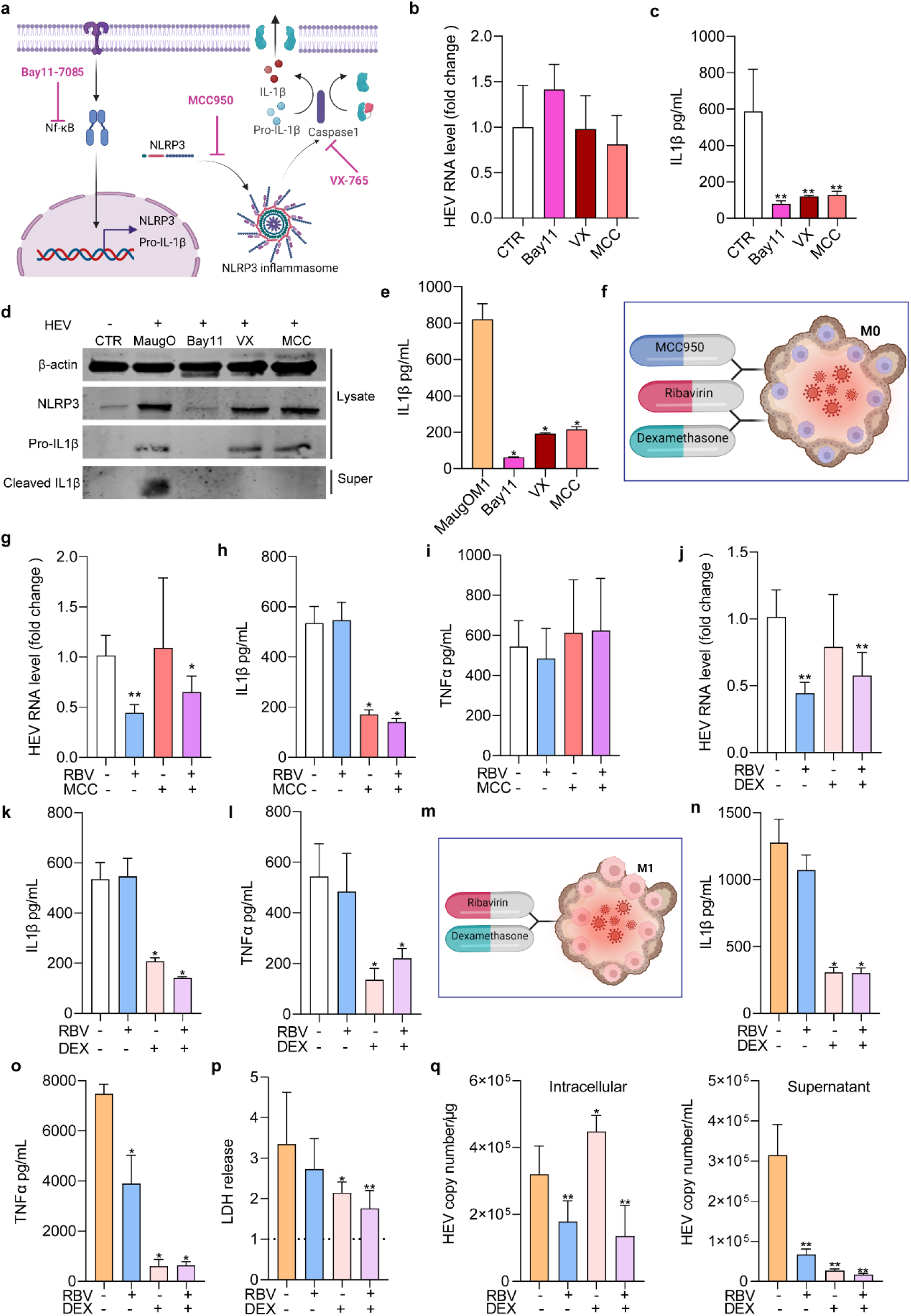
Pharmacological targeting of NLRP3 inflammasome and combination with antiviral treatment in macrophage-augmented organoids. **a**, Schematic illustration of the NLRP3 inflammasome pathway and the pharmacological inhibitors targeting the key components. **b,** The effects of the NF-κB inhibitor (BAY11-7085, Bay11), caspase-1 inhibitor (VX-765;VX) and NLRP3 inhibitor (MCC950; MCC) on HEV RNA level quantified by qRT PCR in infected MaugOs (n = 4). **c**, ELISA quantification of IL-β secretion into supernatant of HEV infected MaugOs (CTR) and infected MaugOs treated with the inhibitors (Bay11, VX or MCC) (n = 4-8). **d,** Western blotting analysis detecting the key components of the NLRP 3 cascade in cell lysate and supernatant (super) of these five groups. **e**, The effects of these three inhibitors on IL-β production in MaugOs integrated with M1 macrophages generated from THP-1 monocytes (n = 4). **f**, Schematic illustration of combination treatment in HEV infected organoids integrated with M0 macrophages. **g-i**, Quantification of HEV viral RNA (n = 4-6),IL-β and TNF-α production in HEV infected MaugOs treated with the antiviral drug ribavirin (RBV) the NLRP 3 inhibitor (MCC) or the combination (n = 4). (**g**) The infected but untreated MaugOs served as control (normalized as 1). **j**-**l**, Quantification of HEV viral RNA (n = 4-6) and protein production of IL-β, TNF-α in HEV infected MaugOs treated with ribavirin, the anti-inflammation drug dexamethasone (DEX) or the combination (n = 4). (j) The infected but untreated MaugOs served as control (normalized as 1). **m**, Schematic illustration of combination treatment in HEV infected organoids integrated with M1 macrophages. **n** and **o**, Production of IL-1β and TNF-α Cytokines in HEV infected MaugOs (M1) treated with ribavirin, dexamethasone (DEX) or the combination (n = 4). **p**, Quantification of lactate dehydrogenase (LDH) release (n = 6). Infected but untreated served as control (normalized as 1, indicated by dash line). **q**, Quantification of intracellular and supernatant HEV RNA levels by qRT-PCR (n = 6). (**a**, **f**, **m**) The schematic illustration was generated in Biorender. (**b**, **c**, **e**, **g**-**l**, **n**-**q**) Data are mean ± SD. *p < 0.05; **p < 0.01; Mann-Whitney U test.

We next tested the combinational treatment in HEV-infected MaugOs integrated with the pro-inflammatory M1 macrophages (Fig. 6m), which has a more prominent feature of inflammatory cell death (Fig. 4i). Potent inhibition on IL-1β (Fig. 6n) and TNF-α (Fig. 6o) cytokine production was observed by dexamethasone or the combination treatment, although not on the level of IL-10 production (Extended Data Fig. 5i). Intriguingly, anti-inflammatory treatment attenuated LDH release, a hallmark of cell death, and this effect was more potent in the combinational treatment group (Fig. 6p). This is in line with the quantification of HEV genome copies that dexamethasone slightly increased viral RNA levels intracellularly, but dramatically inhibited the release of HEV into supernatant, which reflected the inhibition of lytic inflammatory cell death (Fig. 6q). These results collectively demonstrated the advantages of using MaugOs in developing improved treatment strategies aiming at simultaneously targeting viral replication and the resulting inflammatory response, as well as inflammatory cell death.

### MPXV infects human liver-derived organoids

Systemic manifestations appear to be associated with more severe outcomes in mpox patients^7,35^. To probe the potential of hepatic manifestation (Fig. 7a), we analyzed the levels of alanine aminotransferase (ALT), a classical biomarker for liver damage, in a large cohort of mpox patients with advanced HIV infection^36^. Considering the upper limit of normal value for serum ALT as 50 IU/L^37^, among the 203 mpox patents with ALT test available, 42 patients had the values above this threshold. Among these, 37 patients had ALT values between 50 and 200 IU/L, 4 patients were between 200 and 1000 IU/L, but one patient had 7000 IU/L (Fig. 7a). Next, we in-depth examined the liver from a patient died from disseminated mpox^38^, who developed MPXV infection associated hepatitis (Extended Data Fig. 7a-7f). At microscopic exam, we observed various and scattered foci of lobular MPXV-hepatitis, with hypereosinophilic (or acidophilic) hepatocytes, and some presenting Guarnieri-type inclusion in their cytoplasm (Fig. 7b). Immunohistochemical staining identified MPXV infected hepatocytes with enhanced Guarnieri-type paranuclear inclusion or acidophilic degeneration (Figure 7c). Some cholangiocytes showed pyknotic nuclei and eosinophilic cytoplasm (Figure 7d, left panel). Double immunohistochemical staining showed MPXV infection in cholangiocytes, although relatively rare (Figure 7d, right panel).

**Fig. 7:**
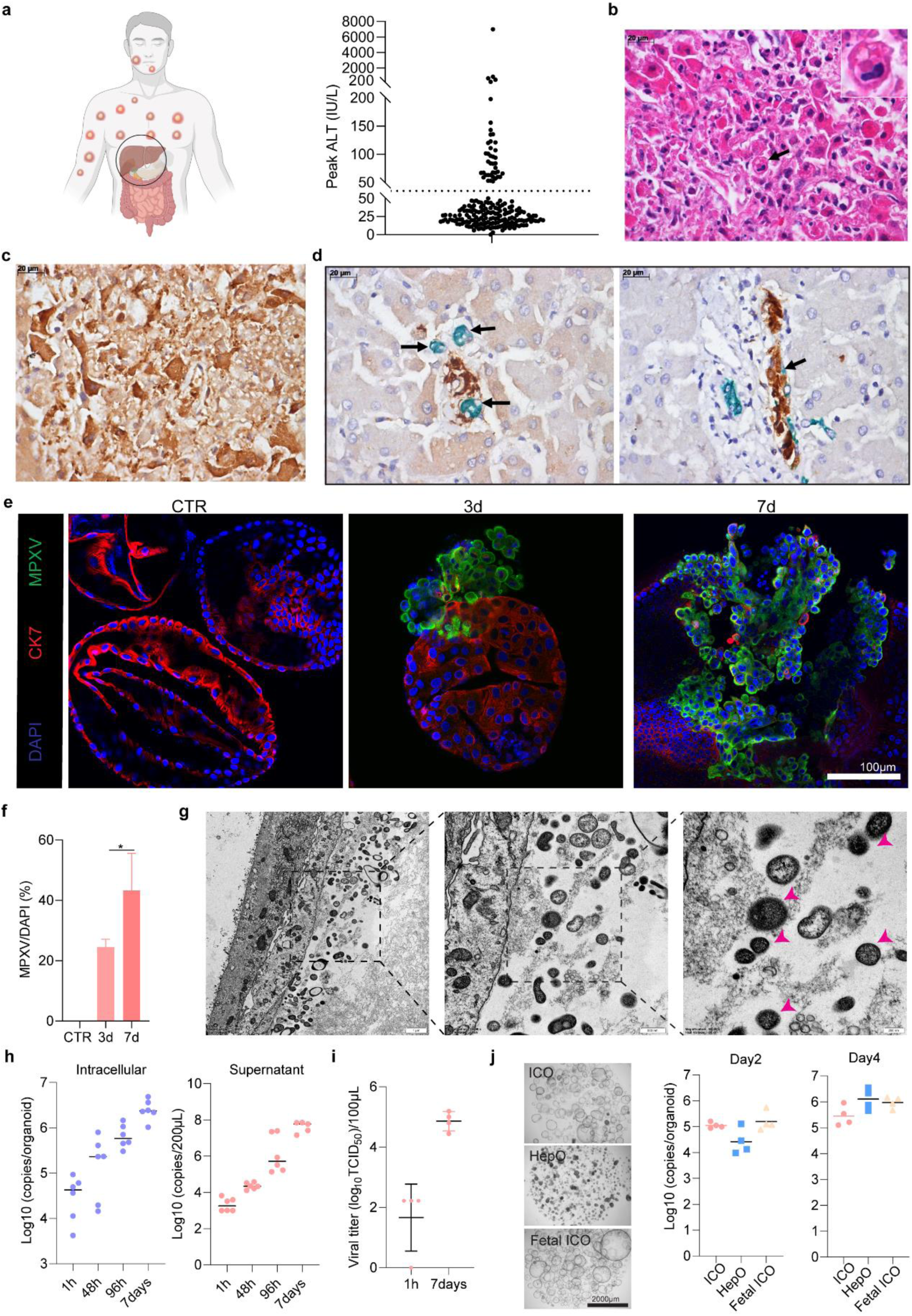
Hepatic manifestations in mpox patients and MPXV infection in human liver-derived organoids. **a**, The Peak ALT values from a cohort of 203 mpox patients. The upper limit of normal valuefor serum ALT (50 IU/L) was indicated by the dash line. **b,** Focal hepatitis, showing eosinophilic degeneration of hepatocytes, some with eosinophilic paranuclear cytoplasmatic inclusion, Guarnieri body type (arrow and inset), hyperplastic Kupffer cells and with scarce inflammatory reaction (H&E staining). **c**, Immunohistochemical staining against the viral antigens visualizes MPXV infected hepatocytes and possibly inflammatory cells. **d**, Double immunohistochemical staining for cholangiocytes (CK7 in green) and for viral antigens (in brown), showing that Biliary ducts are surrounded by MPXV-infected hepatocytes and inflammatory cells (left panel), and some cholangiocytes are also infected by MPXV (right panel). **e**, Confocal imaging of MPXV infection in ICOs derived from adult human liver. CK7 (red): marker of biliary epithelium; MPXV (green): staining against the virions; DAPI (blue): nuclear staining. **f**, Quantification of MPXV-infected cells from confocal images at 3 days post-infection (n = 4) and 7 days post-infection (n = 5). **g**, Electron microscopy visualizing virus particles in ICOs infected with MPXV. The color arrows point to representative MPXV particles. **h**, qRT-PCR quantification of MPXV genomic DNA from infected ICOs and the harvested supernatant at the indicated time points (n = 6). The organoids were counted prior to infection. The number of viral copies per organoid was calculated by dividing the total viral copy number per well by the number of seeded organoids. **i**, Quantification of MPXV virus titers by TCID50 assay in the supernatant harvested at the indicated time points (n = 4). **j**, Representative images of adult human liver-derived ICOs (ICO), hepatic-differentiated ICOs (HepO), and fetal human liver-derived ICOs (Fetal ICO). Quantification of MPXV genomic DNA in these three organoid types at indicated time points post-infection (n = 4). (**f**, **h**, **i**, **j**) Data are mean ± SD. *p < 0.05; **p < 0.01; Mann-Whitney U test.

These clinical observations motivated us to investigate whether human liver-derived organoids are susceptible to MPXV infection. We first tested in ICOs cultured from adult liver. Immunofluorescent staining based on our previously established protocols^39^ for the MPXV virons (Fig. 7e) and viral dsRNA (Extended Data Fig. 7g) demonstrated robust infection. Quantification showed over 20% and 40% MPXV-positive cells after 3 or 7 days infection, respectively (Fig. 7f). The virus particles were also visualized by electron microscopy (Fig. 7g). Quantification of intracellular and secreted genomic viral DNA revealed the increase of viral replication, showing over 2-4 log10 increase in genome copies from 1 hour to day 7 post-inoculation (Fig. 7h). Quantification of virus titer confirmed the infectiousness of produced viruses with over 2 log10 increase in viral titers during this 7-day culture period (Fig. 7i). Next, we comparatively assessed the susceptibility in ICOs, ICO differentiated hepatic organoids, and ICOs cultured from human fetal liver (Fig. 7j). Upon virus inoculation, all three types of organoids robustly supported MPXV infection, shown by quantification of MPXV genome copies in the organoids and supernatant (Fig. 6j and Extended Data Fig. 7h).

### MPXV infection triggers inflammatory response in MaugOs

Immunohistochemical staining of liver section from the mpox patient showed that a number of Kupffer cells (hepatic macrophages) were infected by MPXV, within areas of MPXV-hepatitis or as isolated cells near to areas with hepatitis (Fig. 8a). This triggered us to examine the response of MPXV infection in MaugOs. We constructed MaugOs based on hepatic differentiated and undifferentiated ICOs, which were inoculated with MPXV and integrated with THP-1 macrophages (Fig. 8b-h and Extended Data Fig. 7i-7l). Transmission of MPXV from infected organoids to surrounding macrophages was observed in both cultures (Fig. 8b, 8e and Extended Data Fig. 7i). Significant upregulation of inflammatory gene expression (e.g. IL-1β, IL-6 and TNF-α) and the production of IL-1β (n = 5; p < 0.01) was observed in hepatocyte differentiated organoids based MaugOs upon MPXV infection (Fig. 8c and 8d). However, the activation of inflammatory response appears more profound in ICO-based MaugOs as shown on IL-1β production and the levels of inflammatory gene expression (Fig. 8g and 8h).

**Fig. 8:**
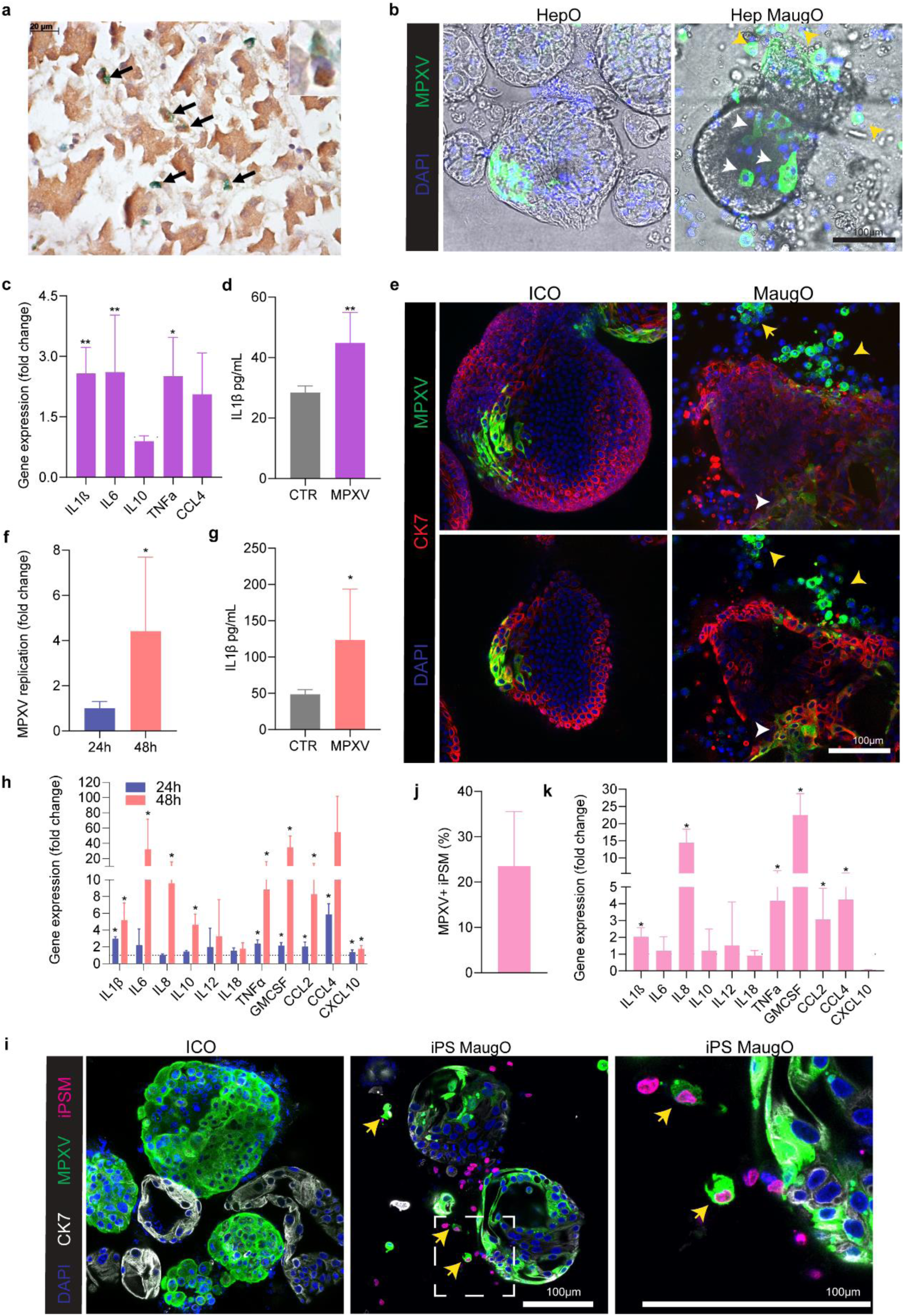
MPXV infection and triggered inflammatory response in macrophage-augmented organoids. **a,** Double immunohistochemical staining for Kupffer cells (CD68 in green) and for viral antigens (in brown), showing MPXV-infected liver macrophages among infected hepatocytes. **b**, Images showing MPXV infection in hepatic differentiated organoids (HepO), and hepatic organoids integrated with THP-1 macrophages (Hep MaugO). White arrows point to MPXV infection possibly in organoids, and yellow arrows point to infection possibly in surrounding macrophages. This was further supported by Extended Data Fig. 7i. **c**, The expression of inflammatory genes quantified by qRT-PCR in MPXV infected Hep MaugOs, compared with the uninfected control (normalized as 1) (n = 5). **d**, Secretion of IL-1β into supernatant in infected (MPXV) and uninfected (CTR) Hep MaugOs, measured by ELISA (n = 5). **e**, 3D constructed (upper) and single layer (lower) images of MPXV infected ICOs (derived from adult human liver) and ICOs integrated with macrophages. White arrows point to infected organoids showing double positive for CK7 and MPXV staining. Yellow arrows point to infected macrophages showing single staining of MPXV. **f**, qRT-PCR quantification of MPXV viral RNA in MaugOs (n = 4). **g**, Secretion of IL-1β into supernatant in infected (MPXV) and uninfected (CTR) MaugOs based on ICOs (n = 4). **h**, The expression of inflammatory genes in MPXV infected compared with uninfected (normalized as 1) MaugOs at 24 and 48 hours (n = 4). **i**, Images showing MPXV infection in ICOs and integrated with hiPSC-derived macrophages (iPS MaugO). Yellow arrows point to infected macrophages. **j**, Quantification of MPXV-infected macrophages (n = 4). **k**, The expression of inflammatory genes quantified by qRT-PCR in MPXV infected iPS MaugOs, compared with the uninfected MaugOs (normalized as 1) (n = 4). (**c**, **d**, **f**, **g**, **h**, **k**) Data are mean ± SD. *p < 0.05; **p < 0.01; Mann-Whitney U test.

Finally, we employed ICO-based MaugOs integrated with a fluorescent protein tagged macrophages generated from hiPSCs for better visualization (Extended Data Fig. 1f-1h). This allowed us to directly visualizing MPXV transmission from infected organoids to macrophages (Fig. 8i), and over 20% of the macrophages are positive for anti-MPXV staining (Fig. 8j). Consistently, this resulted in elevated expression of inflammatory genes (Fig. 8k). Collectively, we demonstrated that human liver-derived organoids with ductal- or hepatocyte-phenotype are susceptible to MPXV infection, and MaugOs simultaneously recapitulate MPXV infection and the resulting inflammatory response.

## Discussion

In respect to modeling viral diseases, the 2D cultured cell lines and even 3D grown human organoids usually only capture the infection life cycle of viruses^39–41^, whereas viral pathogenesis attributes to a magnitude of mechanisms in particular involvement of the host immune system^42^. Research on developing complex experimental models integrating epithelial and immune cell compartments is emerging^43–45^. In this study, we developed macrophage-augmented human organoid systems that enable to simultaneously model the viral life cycle, host inflammatory response, as well as infection triggered cell injury. Here, we focused on viral infections affecting the liver, and prioritized three representative pathogenic viruses—HEV, SARS-CoV-2 and MPXV. The intrahepatic biliary compartment harbors stem cell/progenitor populations that can be cultured into ICOs, which intrinsically carry a cholangiocyte phenotype, but can be differentiated toward a hepatocyte-like phenotype^18,46,47^. Although HEV primarily infects hepatocytes, direct infection of cholangiocytes has been reported in hepatitis E patients who actually developed cholangitis^21^. Consistently, our previous study has demonstrated that human liver-derived organoids regardless cholangiocyte- or hepatocyte-like phenotype can effectively support HEV infection in culture^22^. This study mainly employed ICOs but also hepatic differentiated organoids to build MaugOs for modelling HEV infection and the resulting inflammatory response and cell death.

Since liver manifestations have been rarely investigated during the current mpox outbreak, we performed a post-mortem examination of the liver from a patient who developed MPXV infection associated hepatitis^38^. Immunohistochemical staining showed predominate infection of hepatocytes by MPXV. It can also infect cholangiocytes, but the frequency was very low. Intriguingly, a number of Kupffer cells were infected by MPXV, and bacterial LPS can be detected within the cytoplasm of Kupffer cells. Since this patient had severe intestinal manifestations, we postulate that disruption of the intestinal barrier integrity may lead to bacterial translocation and MPXV spreading to the liver, causing focal hepatitis. In the current outbreak, a case of a 10-day-old neonate with MPXV infection was documented having an elevation of transaminases (ALT and AST; markers for liver injury) on day 6 of admission, which was reduced after treatment with the antiviral drug tecovirmat^48^. In 1987, examination of the liver tissue of a 9-month-old child who died after MPXV infection observed that most of the cells in the assessed sections were infected by the virus, presumably including hepatocytes, cholangiocytes and immune cells^7^. In line with these clinical observations, we found that ICOs derived from fetal or adult human livers, as well as the hepatocyte differentiated ICOs, robustly support productive infection of MPXV. Furthermore, in a large cohort of mpox patients with advanced HIV infection, we found that 20% of the patients (42 out of 203) had elevated ALT levels (above 50 IU/L), although it remains unknown whether MPXV infection is the cause of this elevation. However, this raises the question whether liver function tests should be performed in mpox patients, especially for the subpopulation with proctitis, which has been commonly reported in the current outbreak^49^.

Viruses from infected tissues can spread to macrophages to trigger inflammatory response^50^. This viral spreading process is likely attributed to both active infection and phagocytosis of virus particles or virus-containing cell debris. In our constructed MaugOs, we observed clear evidence that all three tested viruses can be transmitted from infected organoid cells to macrophages, which can replicate in macrophages to trigger inflammatory responses. Genome-wide transcriptomic analysis revealed the activation of key pathways related to infection and inflammation in MaugOs. More interestingly, there is evidence of active interactions between organoids and macrophages, since there are additional pathways that are only activated in MaugOs. In the human liver, macrophage populations are highly heterogeneous, which are further complicated by the types and stages of the diseases^51^. Intriguingly, when HEV infected ICOs integrated with M1 macrophages, inflammatory cell death occurred predominantly in the organoids compartment but also in macrophages, mimicking immune-mediated tissue damage in infected patents. More interestingly, when culturing in a trans-well system separating organoids and macrophages into two compartments, significantly increased levels of cell death were observed in the organoids compartment with HEV infection, whereas HEV infection alone without co-culturing with macrophages has minimal effects on cell death in organoids. These results indicate that HEV triggered inflammatory cell death in MaugOs is at least partially through a paracrine mechanism, but we do not exclude whether direct cell-cell contact is also involved. Nevertheless, productive infection may not be a prerequisite for initiating an inflammatory response in macrophages, at least in the case of HEV. Our previous study has shown that exposure to inactivated HEV particles exerted equivalent potency in activating NLRP3 inflammasome in macrophages compared to that of infectious viruses^32^. As an intuitive comparison of the three viral disease models, HEV appears to provoke the most rapid and potent activation of inflammatory response, and this can be effectively inhibited by pharmacological inhibitor targeting the NLRP3 inflammasome cascade. This is also in line with clinical features of severe acute HEV infection, which can cause massive liver inflammation and acute liver failure especially in pregnant women^52^. Of note, both the basal level and the activation of inflammatory responses by HEV or MPXV are weaker in hepatocyte differentiated organoids integrated with macrophages compared to that of undifferentiated ICOs. We speculate that this could be attributed to the prolonged presence of dexamethasone, a potent anti-inflammatory agent, during hepatic differentiation of the organoids. In the case of SARS-CoV-2, we demonstrated as a proof-of-concept that this prototype model is capable of recapitulating both SARS-CoV-2 replication and the resulting inflammatory response. It could be further developed, for example, into an excellent model for studying post-acute sequelae of COVID-19 (PASC; long COVID), as emerging evidence indicates that specific subsets of PASC are attributed to viral persistence in (extrapulmonary) reservoirs and consequently chronic inflammation^53,54^.

We further explored the use of hepatic (tissue) macrophages isolated from liver graft preservation fluid at the setting of liver transplantation, a unique source of obtaining resident immune cells from the “healthy” human liver^55^. However, we observed the activation of inflammatory genes when hepatic macrophages were integrated into organoids even without viral infection, but this does not occur when integrated with other macrophage types. We postulate this may be attributed to the pre-activation of immune cells during graft perfusion, procurement, or cold storage which are associated with ischemia and tissue injury^55^, although the exact underlying mechanisms require further investigation. We thus developed an alternative approach by establishing a trans-well based co-culture system. We observed the transmission of HEV from infected organoids to macrophages through the paracrine route and robustly activated inflammatory responses.

Bile acids are signal molecules through FXR and TGR5 pathways to regulate lipid, glucose, and energy metabolism^29^. A landmark study has shown that bile is essentially required to support human norovirus replication in intestinal organoids, thus allowing establishment of the first *in vitro* model for human norovirus infection^56^. Bile acids have also been shown to promote the replication of hepatitis B virus and hepatitis C virus^57,58^. In contrast, we here found that human bile and two representative primary bile acids inhibit HEV replication in ICOs. Bile acids have been shown to inhibit the infection of multiple viruses including human immunodeficiency virus^59^, chikungunya virus^60^, and influenza A virus^61^. These studies are not necessarily contradictive, since bile acids can activate multiple signaling pathways mainly through the receptor FXR or TGR5 to exert distinct effects on viral infections^62^. We found that the anti-HEV activity of bile is primarily mediated by FXR, although the detailed mechanisms remain to be further studied. A previous study has shown that bile acids can inhibit LPS-induced expression of pro-inflammatory genes in macrophages, presumably through the TGR5 pathway, although the exact mechanism remains uncertain^63^. Interestingly, we found that bile also inhibit HEV-triggered inflammatory response in MaugOs, which is independent of FXR but at least partially through TGR5. These results demonstrate the strength of our model system in capturing the complex mode-of-actions of human bile on infection and inflammation.

For patients with severe acute infection accompanied with hyperinflammation, antiviral^64,65^ or anti-inflammatory^34,66^ monotherapy may offer some/partial benefits. However, for these patients, we believe the necessity of developing more advanced treatment strategies to simultaneously inhibit viral infection and pathological inflammation. In HEV-infected MaugOs, we demonstrated proof-of-concept of combining antiviral and anti-inflammatory agents to achieve an improved outcome. Ideally, it is more appealing to identify drugs having both antiviral and anti-inflammatory activities. Agents such as niclosamide^67^ and indomethacin^33^ have been shown to possess such properties, but whether their monotherapy would be potent enough remains questionable. Inflammatory cell death has been extensively implicated in severe acute viral diseases in driving tissue damage and patient morbidity and mortality^68^. We found that MaugOs integrated with M1 macrophages recapitulate prominent features of inflammatory cell death upon HEV infection. Adding anti-inflammatory treatment partially, although not completely, inhibited cell death. A recent study has shown that treatment with a pharmacological of inflammatory cell death ameliorated lung inflammation and prevented mortality following influenza A virus infection in mice^69^. We postulate the need of further discovery and development of “multi-actionable” agents or through drug combinations to simultaneously inhibit viral infection, inflammatory response and inflammatory cell death, whereas MaugOs can serve as a unique tool for developing such multitarget therapeutics. Although large-scale drug screening remains challenging in these augmented organoids, we believe it is capable of profiling small- to medium-size (e.g. hundreds to thousands compounds) drug libraries^22^.

Of note, there are some limitations of the study. First, hepatic macrophages obtained from liver graft preservation fluid, but not from healthy human livers, were tested. Although practically and ethically challenging, it would be helpful to further extend the MaugO model by accessing homeostatic macrophages from healthy human livers through other sources whatever possible. Second, other immune cell types (e.g. dendritic cells) as well as stromal cells (e.g. hepatic stellate cells) should be considered to further leverage the model for recapitulating the multi-layered pathophysiology of viral and non-viral diseases. Third, our models remain far from creating an organ *in vitro* or recapitulating the normal tissue anatomy of the liver. Last but not least, this study focused on short-term experiments resembling acute viral diseases. Future research should explore the possibility of studying chronic viral infections in MaugOs, as well as expanding to other pathogenic viruses. Collectively, establishing MaugOs for modeling the complexity of viral diseases shall essentially contribute to the ongoing paradigm shift towards animal-free models for human diseases in biomedical research. Furthermore, these models will advance the development and testing of novel therapeutics.

## METHODS

### Organoids culture

Tissue samples (≤0.5 cm³) from donor liver biopsies, utilized for organoid isolation and culture, were obtained during liver transplantation procedures at the Erasmus Medical Center Rotterdam. The use of liver tissues for research purposes was approved by the Medical Ethical Council of the Erasmus MC, and informed consent was given (MEC-2014-060 for adult liver tissues and MEC2006-202 for fetal liver tissues). Human adult and fetal intrahepatic cholangiocyte organoids (ICOs) were isolated and cultured following previously established procedures.^46^ These ICOs were cultured in organoid expansion medium (EM), formulated with advanced DMEM/F12 (Invitrogen). The EM was supplemented with 100 µg/mL primocin (Life Technologies), 0.01 M Hepes (Life Technologies), 200 mM ultraglutamine (Life Technologies), 1% (v/v) of N2 (Gibco), 2% (v/v) of B27 (Gibco), 1 mM N-acetylcysteine (Sigma-Aldrich), 10 mM nicotinamide (Sigma-Aldrich), 5 μM A83.01 (Tocris), 10 μM forskolin (Tocris), 10 nM gastrin (Sigma-Aldrich), epidermal growth factor (EGF) at 50 ng/mL (PeproTech), 10% (v/v) of R-spondin-1 (conditioned medium), fibroblast growth factor 10 (FGF10) at 100 ng/mL (PeproTech), hepatocyte growth factor (HGF) at 25 ng/mL (PeproTech) (see more details in Supplementary Table S1 and Table S2).

### Hepatocyte differentiation of organoids

When ICOs isolated from adult human liver reaching 75% density in basement matrix (Matrigel Corning), the culture medium EM was switched to EM supplemented with bone morphogenetic protein 7 (BMP7) at a concentration of 25 ng/mL (PeproTech) for a duration of 5 days. At day 5 the BMP7 treated ICOs were harvested, dissociated into smaller fragments and re-seeded within the same environment. The medium was changed to organoid differentiation medium (organoid DM), composed of AdDMEM/F12 medium supplemented with 100 µg/mL primocin (Bio-Connect BV), 1 M Hepes (Life Technologies), 200 mM ultraglutamine (Life Technologies), 1% N2, and 2% B27 (with vitamin A). Additionally, organoid DM contained the following factors: EGF at 50 ng/mL (PeproTech), gastrin at 10 nM (Sigma-Aldrich), HGF at 25 ng/mL (PeproTech), FGF19 at 100 ng/mL (PeproTech), A8301 at 500 nM (Tocris), DAPT at 10 μM (Sigma-Aldrich), BMP7 at 25 ng/mL (PeproTech), dexamethasone at 30 μM (Sigma-Aldrich), IWP2 at 3 μM (Sanbio), iCRT3 (Sigma-Aldrich) at 25 μM, Carbachol (Sigma-Aldrich) at 100 μM, and CHIR99021 (Sigma-Aldrich) at 3 μM. The organoid DM was refreshed every 2 to 3 days, and this differentiation process was continued for a period of 10 days, surpassing a total of 14 days of differentiation.

### Macrophage differentiation

Human monocytic THP-1 cells were cultured in RPMI 1640 medium with 15 ng/mL of phorbol 12-myristate 13-acetate (PMA) at 37°C for 48 hours to generate THP-1 macrophages (M0 macrophages). Followed by PBS washing, cells were treated with 100 ng/mL LPS and 25 ng/mL IFN-γ for M1 macrophage polarization or 25 ng/mL IL-4 and IL-13 for M2 macrophage polarization for an additional 2 days. Cells were washed 3 times with PBS and fresh RPMI 1640 medium was subsequently added. The expression of monocyte and macrophage markers was assessed using flow cytometry. Flow cytometric analysis was conducted on a FACSCanto II flow cytometer (BD Biosciences), and the obtained data were analyzed using FlowJo software (version 10.6.1; Tree Star, Inc.).

### Generation of primary macrophages

Primary macrophages were generated from monocytes following a previously described method^32^. Briefly, peripheral blood mononuclear cells (PBMCs) were isolated from healthy donors (Sanquin, The Netherlands) using Ficoll density gradient centrifugation. Monocytes were then separated from PBMCs through plastic adherence in Iscove’s modified Dulbecco’s medium (IMDM) supplemented with 2% human serum. The isolated monocytes were cultured in IMDM (with ultraglutamine), supplemented with 8% (v/v) inactivated FCS, 1% penicillin/streptomycin, and 50 ng/mL of macrophage colony-stimulating factor, for a period of 7 days to generate mature macrophages.

### Isolation of hepatic macrophages

To minimize the contamination of donor peripheral blood cells, hepatic macrophages were obtained from liver graft perfusates during the second back table flush (Supplementary Table S2). Cells were subjected to Ficoll-Paque density gradient centrifugation and subsequently placed in 48-well plates with RPMI 1640 medium for a 2-hour plastic adherence period. Following this, cells were washed with PBS to remove non-adherent cells, and immunofluorescence staining was performed to conform the expression of the macrophage marker CD68, and undifferentiated THP-1 monocytes and differentiated THP-1 macrophages were set as negative and positive controls, respectively.

### Generation of human induced pluripotent stem cells derived macrophages

This protocol has been detailed described in our previous study.^19^ Briefly, monocytes were first generated from a GFP tagged hiPSC cell line obtained from the Allen Cell Collection. For macrophage differentiation, 0.75 million monocytes were plated onto the FBS coated well and cultured in IMDM/F12 with nine supplements (IF9S), based on IMDM supplemented (Invitrogen) with 50% F12 Nutrient Mix (Invitrogen), 10 ng/mL Polyvinyl alcohol (Sigma), 0.1% Chemically Defined Lipids (Invitrogen), 2% Insulin-Transferrin-Selenium-X (Invitrogen), 40 µL/L Monothioglycerol solution (Sigma), 64 mg/L L-Ascorbic Acid 2-Phosphate (Sigma), 2 mM GlutaMAX (Invitrogen), 1% non-essential amino acids (Invitrogen) and 0.5% Penicillin-Streptomycin. The IF9S medium supplemented 80 ng/mL M-CSF (Sigma) was refreshed every 1 to 2 days, and the differentiation process was continued for a period of 7 days.

### Establishment and characterization of macrophage-augmented organoids

When the organoids reached sufficient confluence (over 75% of basement matrix), they were harvested in cold AdDMEM/F12 medium and subjected to centrifugation at 300 g for 5 minutes at 4°C to remove the Matrigel. Subsequently, the organoids were mechanically dissociated into small fragments. THP-1 macrophages, primary or hepatic macrophages were prepared in advance and then mixed with organoid fragments (cells of organoids: macrophages = around 1:1), respectively. To facilitate cell movement and integration, we tested a series dilutions of Matrigel. Finally, dilution at a ratio of 1:8 with liver expansion medium was found to be optimal. The diluted Matrigel mixture was subsequently pre-coated in wells. Organoids and macrophages were then seeded on the top of the pre-coated, diluted Matrigel. Organoids alone and macrophages alone were cultured under the same conditions as control groups. MaugOs, organoids, and macrophages were treated with 1 μg/mL LPS to assess the function in response to inflammatory stimulus.

To distinguish the cell types, THP-1 macrophages were pre-labeled with 5 µM CFSE staining solution (Thermo fisher scientific, Cat. No. C34554) in PBS at 37°C for 10 minutes. Following this period, the cells were centrifuged and washed with RPMI 1640 medium (10% FCS) to remove the residual dye. Labeled macrophages were subsequently integrated with organoids to form MaugOs. Images of organoids alone, macrophages alone, and MaugOs were visualized by EVOS and confocal microscopy.

### Virus inoculation

For HEV, organoids were fragmented mechanically and exposed to cell culture produced HEV particles (10^^7^ copy numbers/mL) for 6 hours at 37°C. Throughout the inoculation period, organoids were re-suspended every 30 minutes. Following 6-hour inoculation, organoids underwent centrifugation at 300 g for 5 minutes at 4°C, and the supernatant was discarded. Subsequently organoids were thoroughly washed three times with advanced DMEM/F12 to remove residual viruses. Organoids were cultured in Matrigel and incubated at 37°C with 5% CO2 for one day. Organoids without viral infection were cultured at the same conditions and used as non-infected controls. Then, HEV-infected organoids were collected in cold AdDMEM/F12 medium to remove Matrigel. Subsequently, the organoids were mechanically broken down into small fragments to integrate with macrophages, and cultured wells pre-coated with Matrigel. Non-infected organoids integrated with macrophages cultured under the same conditions served as controls.

For SARS-CoV-2, organoids were fragmented into small pieces and exposed to SARS-CoV-2 viral particles at one multiplicity of infection (MOI) for 2 hours. Subsequently, the organoids were cultured in Matrigel for 5 days. Afterwards, organoids were harvested and washed thoroughly with AdDMEM/F12 medium. The organoids were then segmented and integrated with THP-1 macrophages on a 1:8 diluted Matrigel for an additional 2 days.

For MPXV, organoids were broken into small fragments and inoculated with MPXV particles at 0.5 MOI for 2 hours. Following this, the organoids were washed with AdDMEM/F12 medium and cultured in Matrigel for either 2 or 5 days. After 2 days post-infection, organoids were harvested and integrated with THP-1 macrophages for an additional 1 day. Following 5 days post-infection, organoids were harvested and integrated with macrophages for an additional 1 or 2 days.

### Treatment with human bile or bile acids

Briefly, for organoids, HEV subgenomic RNA harboring a luciferase reporter gene or full-length RNA was delivered by electroporation as previously described^22^. Organoids were first collected in cold Opti-MEM, and a mixture of 3 μg HEV genomic RNA and organoids was transferred into a 4-mm electroporation cuvette (CellProjects). After electroporation, the organoids were cultured in Matrigel for 5 days. Culture medium was refreshed with diluted human bile or bile acids and incubated for the indicated duration. The activity of secreted luciferase in the organoid culture medium was measured using QUANTI-Luc™ Gold (InvivoGen). Organoids containing full-length genomic RNA were harvested, and HEV RNA levels were quantified by qRT-PCR. For macrophages, HEV particles and diluted human bile or bile acids were added to the culture medium for the indicated time period. For MaugOs, organoids were first inoculated with HEV particles and then integrated with macrophages. Human bile or bile acids were added for the indicated time period, and HEV RNA levels were quantified by qRT-PCR.

### Immunofluorescence staining and confocal imaging

To preserve the integrity of samples, MaugOs were seeded into μ-Slide 8 Well (ibidi GmbH, 80826) pre-coated with diluted Matrigel. Fixation was performed by exposing the cells to 4% (w/v) paraformaldehyde (PFA) for 10 minutes at room temperature. Subsequently, the sample-containing well plate underwent three washes with PBS, each lasting 5 minutes. Permeabilization of cells was achieved by treating them with PBS containing 0.2% (vol/vol) tritonX100 for 5 minutes, followed by two additional PBS washes lasting 5 minutes each. The plates were then subjected to a blocking step using 1% bovine serum albumin (BSA) in PBS at room temperature for 1 hour. Next, primary antibodies diluted in 1% BSA were incubated with samples at 4°C overnight. Primary antibodies used in this study are as follows: anti-HEV ORF2 antibody (1:250, mouse), anti-EpCAM (1:500, rabbit), anti-dsRNA antibody (1:500, mouse), anti-CK7 (1:300, mouse), ORF2-specific rabbit hyperimmune serum (1:1000, rabbit; gifted by Prof. Rainer Ulrich, Friedrich Loeffler Institute, Germany), anti-SARS-CoV-2-nucleocapsid protein (1:500, rabbit), anti-CD68 antibody (1:200, mouse), anti-Vaccinia virus Lister Strain (1:1000, rabbit), anti-albumin (1:500, rabbit). Incubation with secondary antibodies took place at room temperature for 1 hour. Next, nuclei were counterstained with DAPI (4,6-diamidino-2-phenylindole; Invitrogen). Subsequently, fluorescence imaging was conducted using a Leica SP5 confocal microscope with a 40× oil immersion objective or a Leica 20× water dipping lens on Leica DM6000 CFS microscope to analyze the stained cellular structures.

### Time-lapse microscopy

The organoids fragments from ICOs and CFSE pre-labeled THP-1 macrophages were cultured in a glass-bottom 96-well plates (SensoPlate 24 well, Greiner Bio-One). The cell cultures were subjected to imaging using bright-field microscopy, employing the sophisticated Opera Phenix system by PerkinElmer, which was equipped with a 10×air objective (NA 0.3). To capture the dynamic process of MaugOs formation, a small z stack was acquired at 30-minute intervals over a 24-hour period, ensuring optimal optical focus at multiple points throughout the experiment.

### Hematoxylin and Eosin (H&E) staining

MaugOs were subjected fixation in 2% paraformaldehyde for 20 minutes, followed by paraffin embedding and sectioning into 4 μm-thick slides. Subsequently, H&E staining was conducted in accordance with standard protocols.

### Genome-wide RNA sequencing and data analysis

HEV infected organoids were integrated THP-1 macrophages for 24 hours to generate HEV-infected MaugOs. THP-1 macrophages were inoculated with HEV particles for 24 hours. Organoids were inoculated with HEV. In parallel, non-infected organoids, macrophages, and MaugOs were cultured under same conditions and served as negative controls. For RNA extraction, total RNA was isolated using the Macherey-Nagel NucleoSpin RNA II Kit (Bioke, Netherlands). The extracted RNA was initial quantified by using the Bioanalyzer RNA 6000 Picochip. Subsequently, RNA sequencing was conducted by Novogene, employing a paired-end 150 bp (PE 150) sequencing strategy.

Correlation of the gene expression levels between samples was calculated according to all gene expression levels of each sample. The correlation coefficient of samples between groups is drawn as heat maps/matrix. The closer the correlation coefficient is to 1, the higher similarity the samples have. The interactions of multiple genes may be involved in certain biological functions. KEGG (Kyoto Encyclopedia of Genes and Genomes) pathway analysis was performed based on differentially expressed genes. KEGG pathways with padj < 0.05 are considered significant enrichment. RNA sequencing data are publicly available at Data Archiving and Networked Services (DANS) https://doi.org/10.17026/LS/W57FUZ.

### Trans-well co-culture of organoids and hepatic macrophages

After a 6-hour inoculation with HEV particles, organoids were cultured in Matrigel in the lower compartment of the trans-well system. Hepatic macrophages were seeded onto 0.4 µm semipermeable transwell inserts (Corning BV) for 24 and 48 hours at a 1:1 ratio with organoid cells, which had been pre-coated with Matrigel. Control groups underwent the same procedural steps without exposure to viral particles. RNA samples were collected from both macrophages in the insert and organoids in the lower compartment simultaneously. The hepatic macrophages from the inserts were collected for CD68 and HEV ORF2 immunofluorescence staining.

### Quantification of HEV genome copy numbers

The quantification of HEV genome copy numbers followed established protocols^22^. Briefly, plasmids harboring the full-length HEV genome (Kernow-C1 p6 clone; GenBank Accession Number JQ679013) served as templates for quantifying HEV genome copy numbers. To create a standard curve, a range of plasmid dilutions, spanning from 10^^-1^ to 10^^-9^, were prepared. Subsequently, these dilutions were amplified and quantified by quantitative reverse transcription polymerase chain reaction (qRT-PCR) to generate a standard curve by plotting the logarithm of the copy number against the cycle threshold (CT) value.

### HEV re-infection assay

HEV-infected organoids were integrated with M0, M1, and M2 macrophages for one day. Supernatants were then collected to inoculate human liver Huh7 cells for 6 hours. Subsequently, the cells were washed for three times with 1 × PBS to remove residual viruses. Following the washes, the cells were incubated with culture medium at 37°C with 5% CO2. The infectivity was assessed through qRT-PCR of viral genome and confocal imaging of viral protein ORF2 expression, respectively.

### Enzyme-linked immunosorbent assay (ELISA)

The concentrations of interleukin-1 beta (IL-1β) in the culture supernatant of macrophage-augmented organoids were determined using an Enzyme-Linked Immunosorbent Assay (ELISA) Kit from BD Biosciences. The concentrations of interleukin-6 (IL-6), interleukin-10 (IL-10) and tumour necrosis factor alpha (TNF-α) were measured by the IL-6 Kit (ThermoFisher Scientific, Cat# 88-7066-22), IL-10 Kit (ThermoFisher Scientific, Cat # 88-7106-88) and TNF-α Kit (ThermoFisher Scientific, Cat # 88-7346-88), respectively.

### LDH release measurement

The release of lactate dehydrogenase (LDH) into cell culture supernatant was assessed using the CytoTox 96® Non-Radioactive Cytotoxicity Assay Kit (Promega), following the manufacturer’s recommended protocols. In brief, the supernatant of HEV-infected organoids integrated with M0, M1, or M2 macrophages derived from THP-1 cells for 24 hours was collected. The LDH activity in the culture supernatant was then measured at a 490 nm wavelength.

### AlamarBlue assay

Culture supernatant was removed, and the organoids or macrophages were incubated with a 1:20 dilution of AlamarBlue (Invitrogen, DAL1100) in culture medium for 2 hours at 37°C. Subsequently, the medium was collected for analysis metabolic activity. Absorbance measurements were obtained using a fluorescence plate reader (CytoFluor Series 4000, PerSeptive Biosystems) at an excitation wavelength of 530/25 nm and an emission wavelength of 590/35 nm.

### TCID50 assay

Viruses in the supernatant of MPXV-infected organoids were harvested, and virus titers were quantified using a 50% tissue culture infectious dose (TCID50) assay. In brief, ten-fold dilutions of the supernatant were inoculated onto vero-E6 cells, which were seeded in a 96-well tissue culture plate at 2,000 cells per well. The plate was incubated for 7 days, and each well was observed under a light microscope for cytopathic effects (CPE). The TCID50 value was calculated using the Reed-Muench method.

### DNA extraction and quantification of MPXV genome copy numbers

Total DNA was purified from MPXV infected organoids or supernatant using Macherey-Nagel NucleoSpin DNA Kit (Bioke, Netherlands). MPXV copy numbers were determined following an established protocol.^39^ Briefly, viral DNA was extracted from purified MPXV, serving as a template for quantifying the genome copy number. Ten-fold serial dilutions of viral DNA ranging from 10^ ^0^ to 10^ ^-7^were prepared and subsequently quantified by qRT-PCR to generate a standard curve. This standard curve was created by plotting the log copy number against the CT value.

### qRT-PCR quantification of gene expression

Total RNA was extracted using the Macherey-Nagel NucleoSpin RNA II Kit (Bioke, Leiden, Netherlands). The concentration and purity were measured using the Nanodrop ND-1000 (Wilmington, DE, USA). Gene expression levels were quantified by SYBR Green–based qRT-PCR using the Applied Biosystems SYBR Green PCR Master Mix (Thermo Fisher Scientific Life Sciences) with the StepOnePlus System (Thermo Fisher Scientific Life Sciences). The housekeeping gene used for normalization was glyceraldehyde 3-phosphate dehydrogenase (GAPDH). Relative gene expression was normalized to GAPDH using the 2^−ΔΔCT method (ΔΔCT = ΔCT_sample − ΔCT_control). Each qRT-PCR experiment included template control and reverse transcriptase control. All primer sequences were listed in Supplementary Table S3.

### Immunoblot analysis

Sample lysates were heated at 95°C for 5 minutes. Proteins were subjected to a 10% sodium dodecyl sulfate polyacrylamide gel (SDS-PAGE), separated at 90 V for 120 minutes, and electrophoretically transferred onto a PVDF membrane (pore size: 0.45 μm; Thermo Fisher Scientific Life Sciences) for 120 minutes with an electric current of 250 mA. Following the transfer, the membrane was blocked with blocking buffer (Li-Cor Biosciences). Primary antibodies were applied and allowed to incubate overnight at 4°C. Primary antibodies used in the study are as followes: anti-NLRP3 antibody (1:1000, Rabbit), anti-IL-1β (1:1000, rabbit), anti-Cleaved-IL-1β (1:1000, rabbit), anti-Cleaved-caspase3 (1:1000, Rabbit), anti-β-actin (1:1000, mouse). Subsequently, the membrane underwent three washes, followed by a 1-hour incubation with anti-rabbit or anti-mouse IRDye-conjugated secondary antibodies (1:5000; Li-Cor Biosciences) at room temperature. After three additional washes, protein bands were visualized using Odyssey 3.0 software.

### H&E and immunohistochemical staining of liver sections from mpox patients

Two patients with confirmed MPXV infection were referred to the central morgue of the city of São Paulo (SVOC-USP). The study was approved by HC-FMUSP Ethical Committee (protocol no. 3951.904). Autopsies were performed at the SVOC-USP after written consent from first-degree relatives^38^. Tissue samples were fixed in buffered 10% formalin for 48 hours, embedded in paraffin and stained with hematoxylin and eosin (H&E) and other special stains. Immunohistochemistry (IH) procedures were performed to detect antigens of MPXV, CD68 for macrophages, CK7 for cholangoicytes, E-cadherin for epithelial adhesion molecules among hepatocytes and LPS for Gram-negative bacterial translocation. Double staining was performed to detect MPXV viral antigen(s), CK7 and CD68. Paraffin sections mounted onto 3-amino-propyltriethoxysilane (cod. A3648, Sigma Chemical, St Louis, MO, USA)-coated glass slides were deparaffinized in xylene, rehydrated, and rinsed in phosphate-buffered saline (PBS), pH 7.4. For antigen retrieval, sections were treated in a 10 mM citrate buffer (pH 6.0). Endogenous peroxidase was inactivated by covering the sections with 3% H2O2. Sections were then incubated with the antibody anti-Vaccinia virus (Thermo Fisher Scientific, PA1-7258) diluted at 1:1500 in a humidified chamber at 37°C for 1 hour. We used UltraVision LP Detection System, polymer –horseradish peroxidase (HRP) (Thermo Fisher Scientific, UK ref. TL-125-HL) staining was completed using the chromogen solution, 3,3’-diamino-benzidin (DAB). After protein block serum-free blockade, the second polyclonal antiserum against CK7 or CD68 was applied and incubated at 4°C overnight. Again the same amplification system was used. The color development (green) was achieved with Perma Green (Diagnostic BioSystems – USA cat.# K074).

### Analysis of ALT values from a mpox cohort

ALT values were obtained from a previous described large cohort of mpox patients with HIV and low CD4 cell counts (CD4 <350 cells per mm3)^36^. Data were provided as de-identified individual level values.

## Statistics

Statistical analysis was performed using the non-paired, non-parametrictest (Mann– Whitney U test; GraphPad Prism software). All results were presented as mean ± SD (p <0.05*; P <0.01**, P <0.001***, p < 0.0001****).

## Data Availability

All data produced in the present study are available upon reasonable request to the authors

## Acknowledgments

Part of this work is supported by the COVID-19 Programme (No. 50-56300-98-2201 to Q.P.) and a VIDI grant (No. 91719300 to Q.P.) from the Netherlands Organisation for Health Research and Development (ZonMw). We thank the Allen Cell Collection for providing the H2B-GFP hiPSC line.

## Author contributions

Conceptualization: Q.P., L.J.W.L., and K.L.; methodology, visualization, and data curation: K.L., Yining W., J.L., J.Z., A.M.G., Z.D., R.S., K.O.V., P.P.C.B., D.M.O. F.H., T.T. and M.E.R.; formal analysis, and investigation: M.M.A.V., Yijin W., H.L.A.J., I.A., C.S., M.P.P., C.M.O., P.L., O.M., A.N.D.N., V.V.O., L.J.W.L., and Q.P.; project administration: K.L., and Q.P.; supervision: Q.P., and L.J.W.L.; writing: Q.P., K.L. and A.N.D.N.; funding acquisition: Q.P.

## Declaration of interests

The authors have no conflict of interests to declare.

## Extended Data Figures & Legends

**Extended Data Fig. 1:**
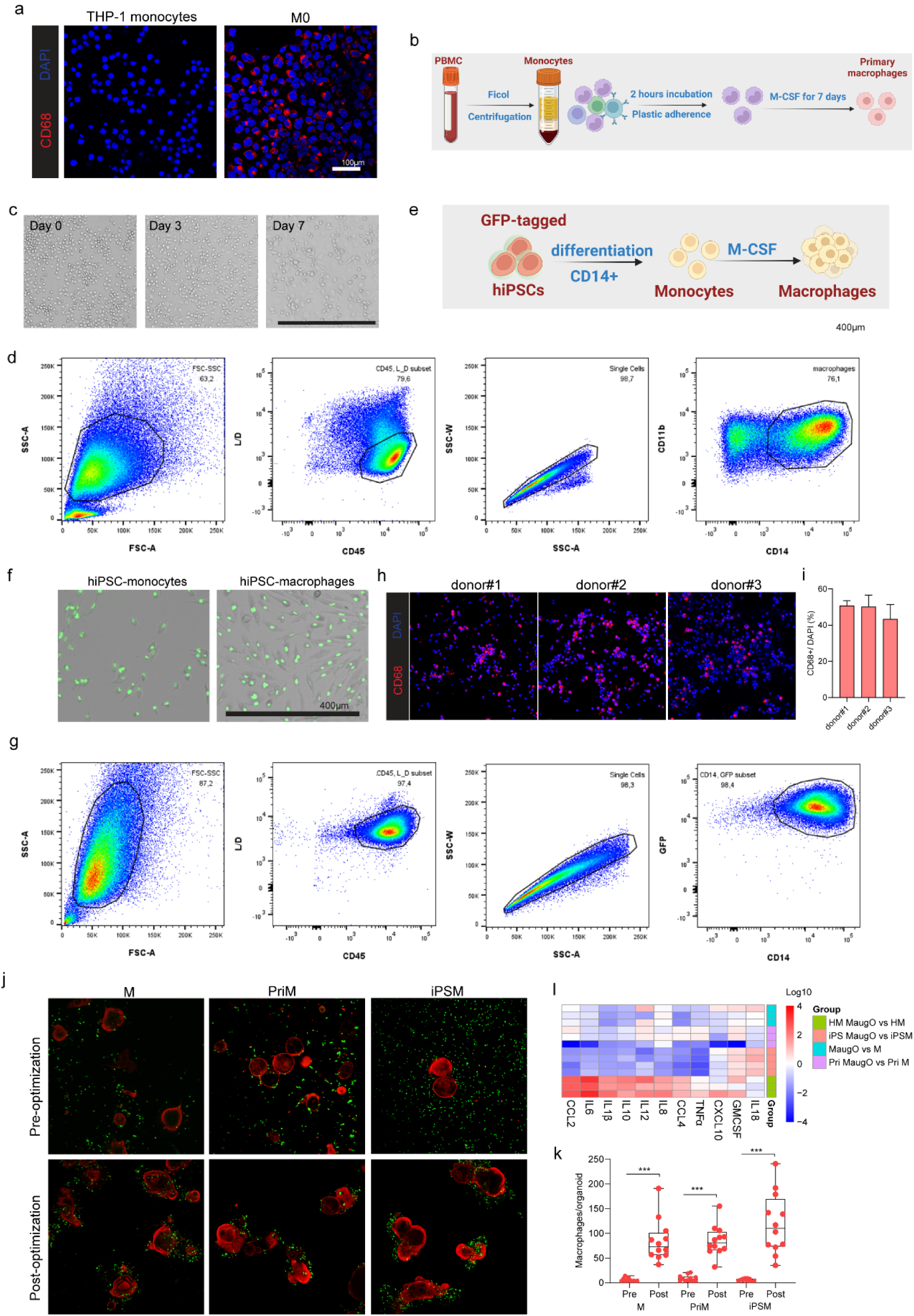
Construction and characterization of macrophage-augmented organoids. a, Immunofluorescence staining images of THP-1 monocytes and differentiated macrophages. CD68, a macrophage marker; DAPI, nuclei. b and c, Schematic illustration of isolation and representative images of primary macrophages during the differentiation process. d, Characterization of primary macrophages by the monocyte/macrophage markers CD11b and CD14. e, Schematic overview of macrophage differentiation from GFP-labeled hiPSCs. f, Brightfield images of hiPSC-derived monocytes and macrophages. g, Characterization of hiPSC-derived macrophages by the monocyte/macrophage marker CD14. h and i, Representative immunofluorescence images and quantification of hepatic macrophages (CD68) from different donors (Donor 1, n = 4; Donor 2, n = 4; Donor 3, n = 2). j and k, Immunofluorescence staining and quantification of individual organoids (from the same line) incorporated with THP-1 macrophages (M; labeled with CFSE; green), PBMC-derived primary macrophages (PriM; stained with CD68; green), and iPSC-derived macrophages (iPSM; tagged with GFP; green). Organoids were stained with EpCAM (red). Statistical analysis comparing pre- and post-optimized protocols was performed by Mann–Whitney U test. n = 12; ***P <0.001. l, Gene expression analysis of MaugOs compared to their corresponding macrophage types: HM MaugO compared to hepatic macrophages (three donors), MaugO compared to THP-1 macrophages (n = 3), and iPS MaugO compared to iPSC-derived macrophages (n = 4). All gene expression thresholds were set to 1. (b and e) The schematic illustration was generated in Biorender.

**Extended Data Fig. 2:**
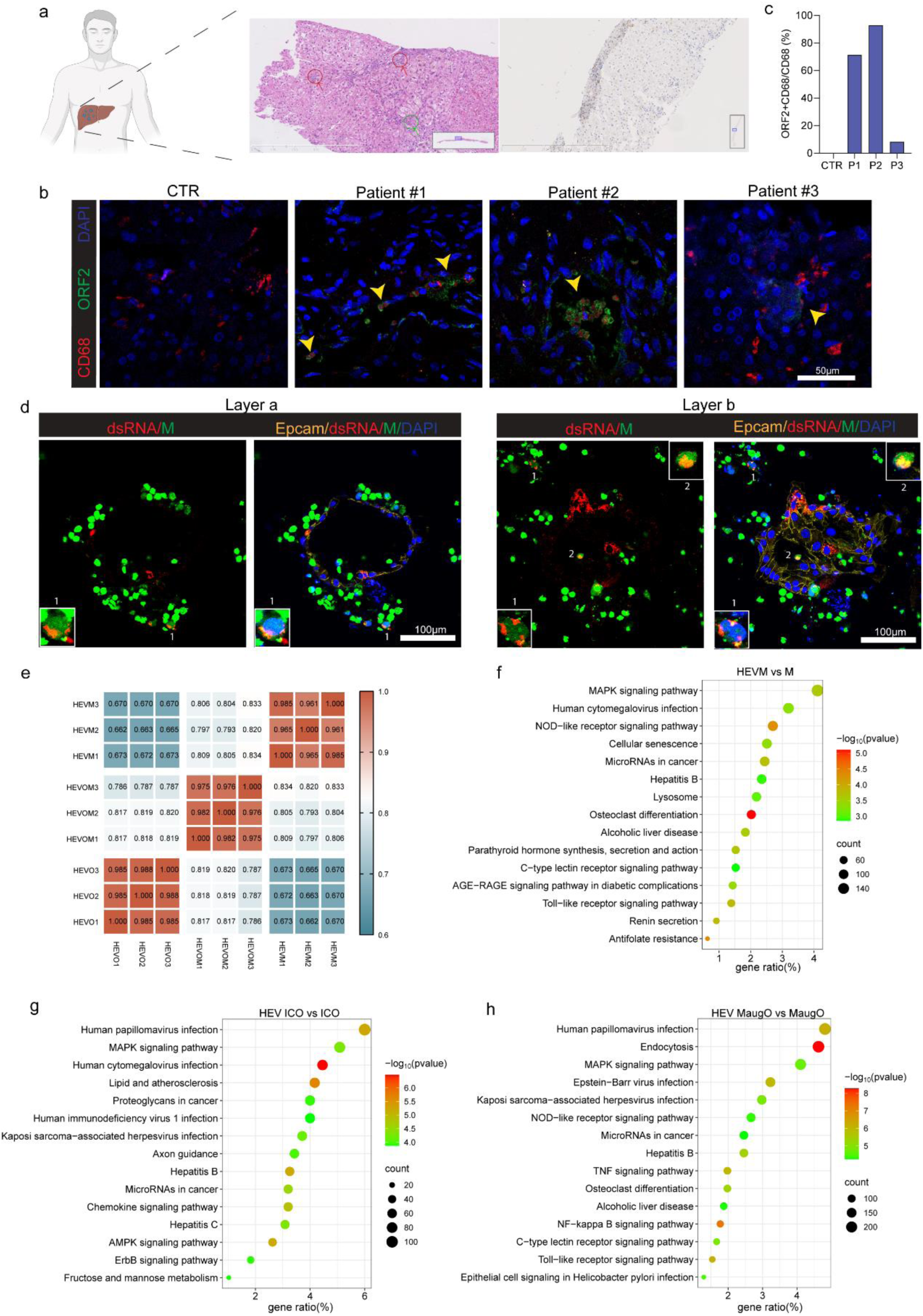
HEV infection in patient liver and responses to HEV infection in macrophages, organoids, and MaugOs. a, Histology and pathology analysis in the liver tissue of a patient with HEV infection. Red circles indicate potentially affected hepatocytes and infiltrated macrophages, and green circle indicates possible involvement of cholangiocytes alone with infiltrated macrophages (left). Immunohistochemical staining of HEV ORF2 capsid protein (right). b, Immunofluorescence staining to detect possible HEV infection in macrophages in liver tissues from three hepatitis E patients. Liver tissue from a patient without HEV infection served as a control. Macrophage marker CD68 (red), HEV ORF2 capsid protein (green), nuclear staining (blue). c, Quantification of HEV ORF2 and CD68 double-positive cells within the CD68 population in patient liver tissues. d, Confocal imaging of viral double-stranded RNA (dsRNA) in HEV infected organoids integrated with THP-1 macrophages for 48 hours. Images from the same view but different layer of Fig. 3b. e, Correlation matrix of transcriptomic profiles of HEV infected organoids, macrophages and MaugOs (HEVO, HEVM, and HEVOM) (n = 3). A value of 1 represents complete correlation, and a value of 0 represents no significant correlation. f-h, KEGG pathway analysis showing top 14 significantly regulated pathways for HEV infected macrophages (M), organoids (ICO), and MaugOs in comparison with their respective uninfected control groups (n = 3). Commentary to Fig. 3e.

**Extended Data Fig. 3:**
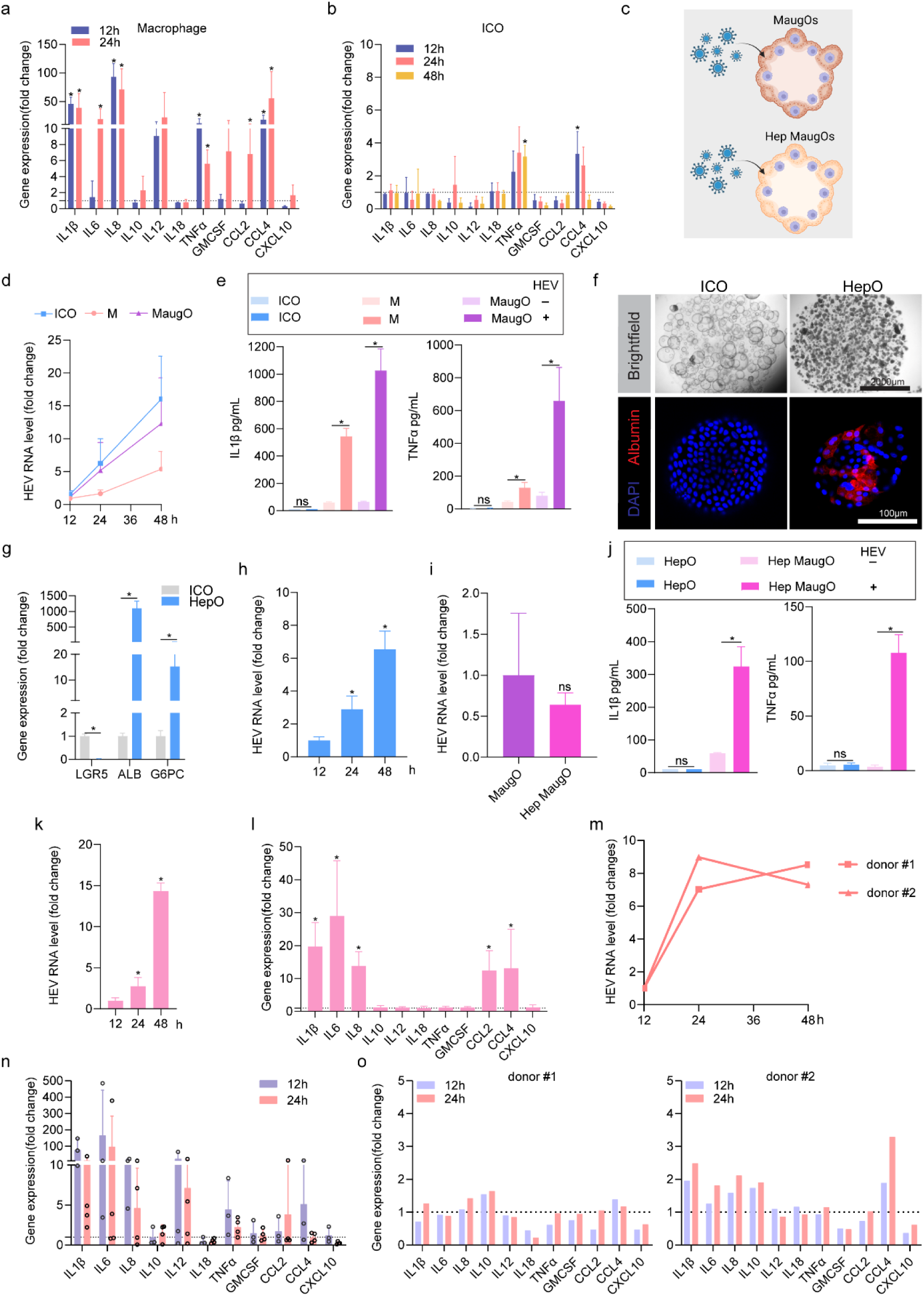
Responses to HEV infection in macrophages, organoids, and MaugOs. a and b, Quantification of inflammatory gene expression in THP-1 macrophages (n = 4) or organoids (n = 4) upon HEV infection for indicated time points. Commentary to Fig. 3e. c, Schematic illustration of experimental design. Cholangiocyte organoids or hepatic differentiated organoids were first integrated with THP-1 macrophages for 24 hours to form MaugOs or Hep MaugOs, and then inoculated with HEV particles. The schematic illustration was generated in Biorender. d, HEV infection kinetics in organoids, macrophages, or MaugOs (n = 6). e, Quantification of IL-1β and TNF-α cytokines in supernatant of organoids (ICO), macrophages (M), or MaugOs (n = 4). f, Brightfield and immunofluoresent imaging of hepatic differentiated organoids. Albumin (red): hepatocyte marker; DAPI (blue): nuclear staining. g, qRT-PCR quantification of the expression of stem cell marker (LGR5) and hepatic markers (albumin/ALB and glucose-6-phosphatase catalytic subunit/G6PC) in undifferentiated (ICO) and hepatic differentiated ICOs (HepO) (n = 4). h, Relative levels of HEV RNA (12h normalized as 1) in hepatic differentiated organoids upon infection (n = 4). i, Quantification of HEV RNA level in MaugOs and Hep MaugOs 24 hours after HEV infection for 24 hours (n = 4). j, Production of IL-1β and TNF-α cytokines in supernatant of hepatic differentiated organoids (HepO), and Hep MaugOs upon HEV infection for 24 hours (n = 4). k and l, Relative levels of HEV RNA (12h normalized as 1) and the expression of inflammatory genes quantified by qRT-PCR in HEV infected hiPSC-derived macrophages (n = 4). m, Quantification of HEV viral RNA in hepatic macrophages derived from perfusates of two donors inoculated with HEV for indicated time points (n = 4). Hepatic macrophages inoculated with HEV for 12 hours set as control (normalized as 1; each donor normalized to the corresponding control). n, Quantification of inflammatory gene expression in hepatic macrophages inoculated with HEV for 12 hours (three donors) and 24 hours (four donors). Uninfected groups served as control (normalized as 1). o, Quantification of inflammatory gene expression in HEV infected organoids integrated with hepatic macrophages (from two donors) for 12 or 24 hours. Uninfected MaugOs served as control (normalized as 1). (a, b, e, g, h, i, j k and l) Data are mean ± SD. *p < 0.05; **p < 0.01; Mann-Whitney U test.

**Extended Data Fig. 4:**
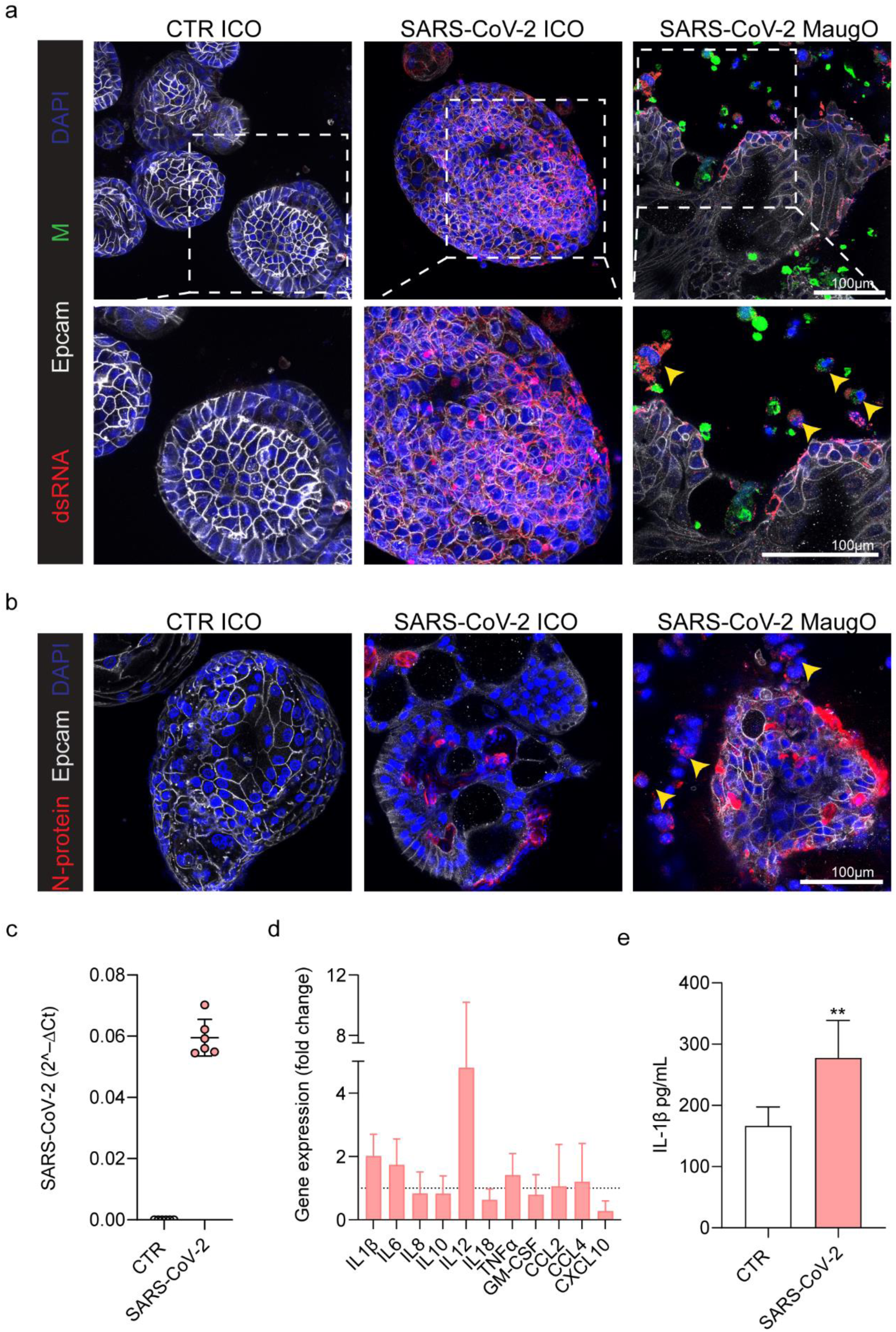
SARS-CoV-2 infection and inflammatory response in macrophage-augmented organoids, related to Figure 2. a and b, Confocal imaging of SARS-CoV-2 infected organoids(SARS-CoV-2 ICO) and SARS-CoV-2 infected organoids integrated with THP-1 macrophages (SARS-CoV-2 MaugO). Yellow arrows point to SARS-CoV-2 transmission from infected organoids to macrophages based on positive for viral dsRNA or N-protein. Macrophages were pre-labeled by CFSE (green) in a. EpCAM marks epithelial (organoid) cells. C, Quantification of SARS-CoV-2 RNA level in infected organoids integrated with THP-1 macrophages for 48 hours. The MaugOs without infection served as control (CTR). d and e, Quantification of inflammatory gene expression (n = 6) and IL-β secretion (n = 6) into supernatant in SARS-CoV2 infected organoids integrated with THP-1 macrophages for 48 hours. Uninfected MaugOs served as control (CTR), and normalized as 1 in panel d. (d, e) Data are mean ± SD. *p < 0.05; **p < 0.01; Mann-Whitney U test.

**Extended Data Fig. 5:**
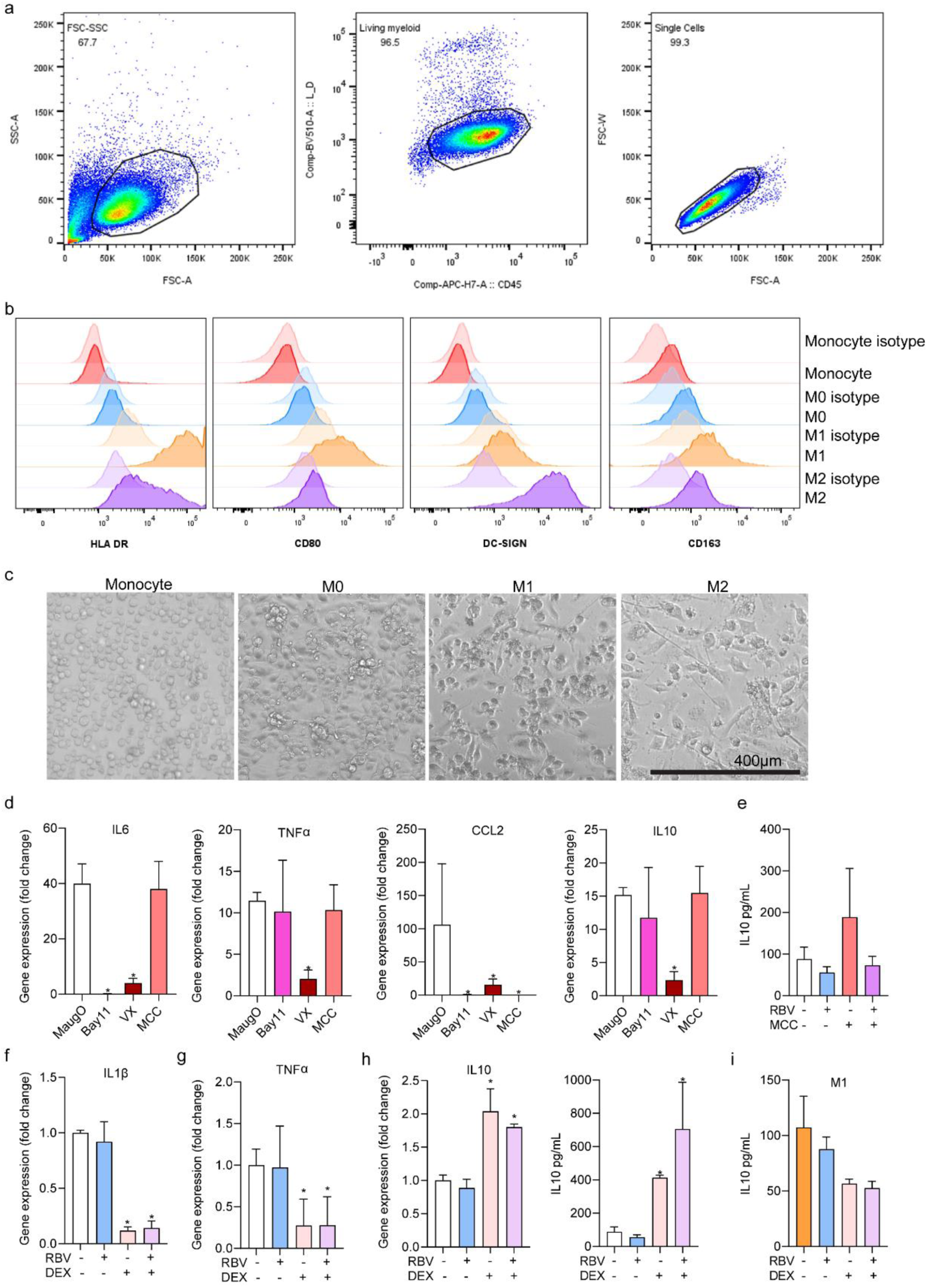
Characterizing macrophages and the response to treatment. a, Gating strategies for flow cytometry analysis of macrophage markers. b, Expression of M1 markers (HLA DR, CD80) and M2 markers (DC-SIGN, CD163) in undifferentiated THP-1 monocytes, and M0, M1, M2 macrophages derived from THP-1 monocytes. c, Representative images of macrophage subtypes. d, The effects of the NF-κB inhibitor (BAY11-7085, Bay11), caspae-1 inhibitor (VX- 765;VX) and NLRP3 inhibitor (MCC950; MCC) on IL-6, TNF-α, CCL2 and IL-10 RNA levels quantified by qRT-PCR in HEV infected MaugOs (n = 4), complementary to Fig. 6c. e, IL-10 cytokine production quantified by ELISA (n = 4),), complementary to Fig. 6i. f and g, Quantification of IL-1β and TNF-α gene expression in HEV infected MaugOs treated with ribavirin (RBV), dexamethasone (DEX) or the combination (n = 4), complementary to Fig. 6k and 6l. h, Quantification of IL-10 RNA and protein levels (n = 4). i, IL-10 cytokine production in HEV infected MaugOs (M1) treated with ribavirin, dexamethasone (DEX) or the combination (n = 4), complementary to Fig. 6o. (d-i) Data are mean ± SD. *p < 0.05; **p < 0.01; Mann-Whitney U test.

**Extended Data Fig. 6.**
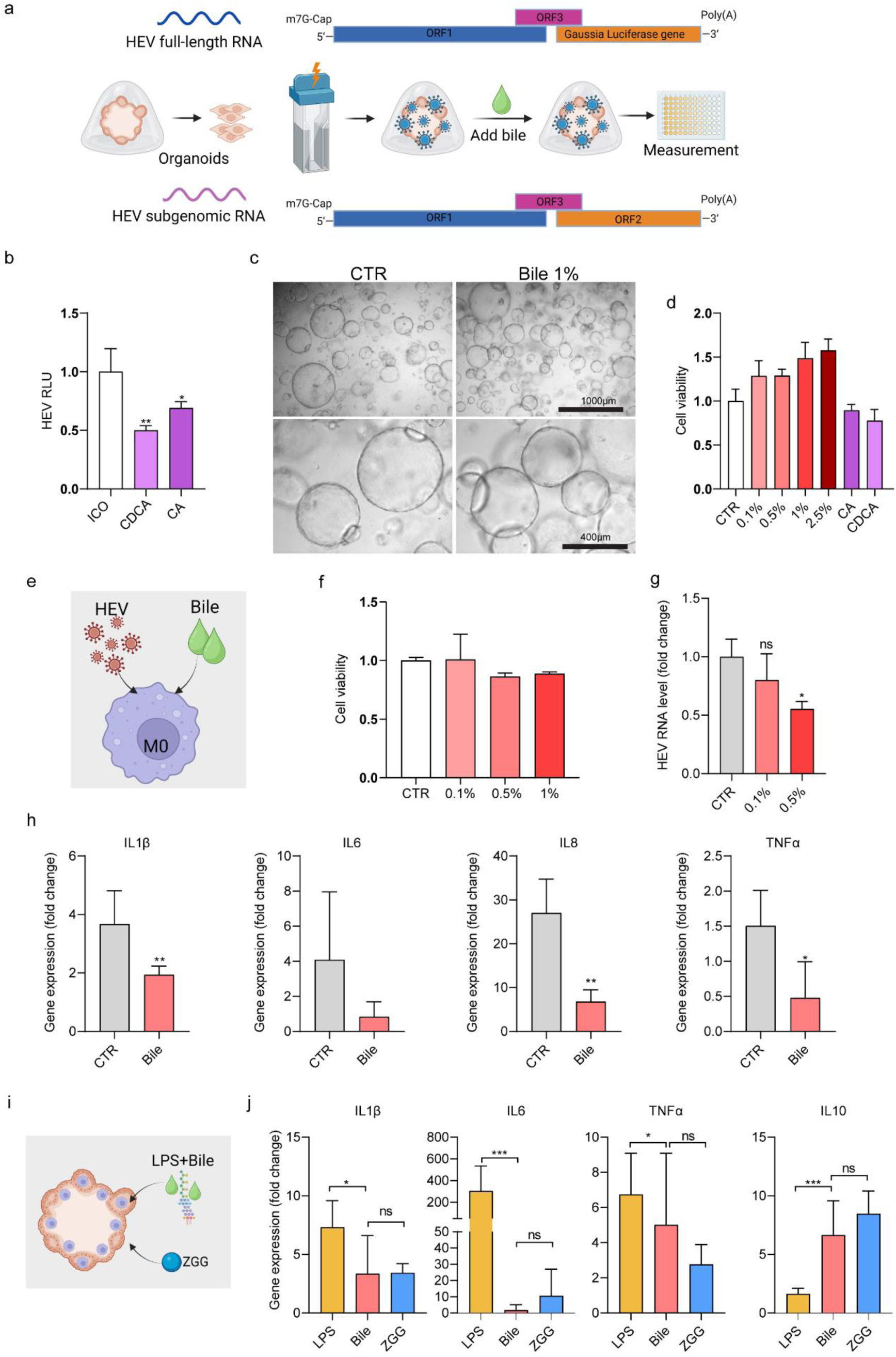
Human bile on HEV infection and inflammatory response. a, Schematic illustration of HEV subgenomic replication model and HEV full-length infectious model with bile treatment. b, The effects of bile acids (100 μM CDCA or 100 μM CA) treatment for two days on organoids (ICO) harboring the HEV subgenomic replicon (n = 6), complementary to Fig. 5d. c, Brightfield images of 1% bile treatment for two days on organoids. d, Cell viability of organoids treated with indicated concentrations of human bile for two days (n = 6). e, Schematic illustration of bile treatment on HEV infected THP-1 macrophages. f, Cell viability of THP-1 macrophages treated with indicated concentrations of bile for one day (n = 6). g and h, Quantification of HEV viral RNA (n = 4) and inflammatory gene expression in HEV infected macrophages treated with 1% bile for one day (n = 4-6). i and j, Quantification of gene expression of IL-β, IL-6, TNF-α and IL-10 in 1 μg/mL LPS incubated MaugOs treated with bile or co-treated with ZGG for one day (n = 4-8), complementary to Fig. 5k and 5l. Data are mean ± SD. *p < 0.05; **p < 0.01; ***p < 0.01; Mann-Whitney U test. (a, e and i) Schematic illustration was generated by biorender.

**Extended Data Fig. 7:**
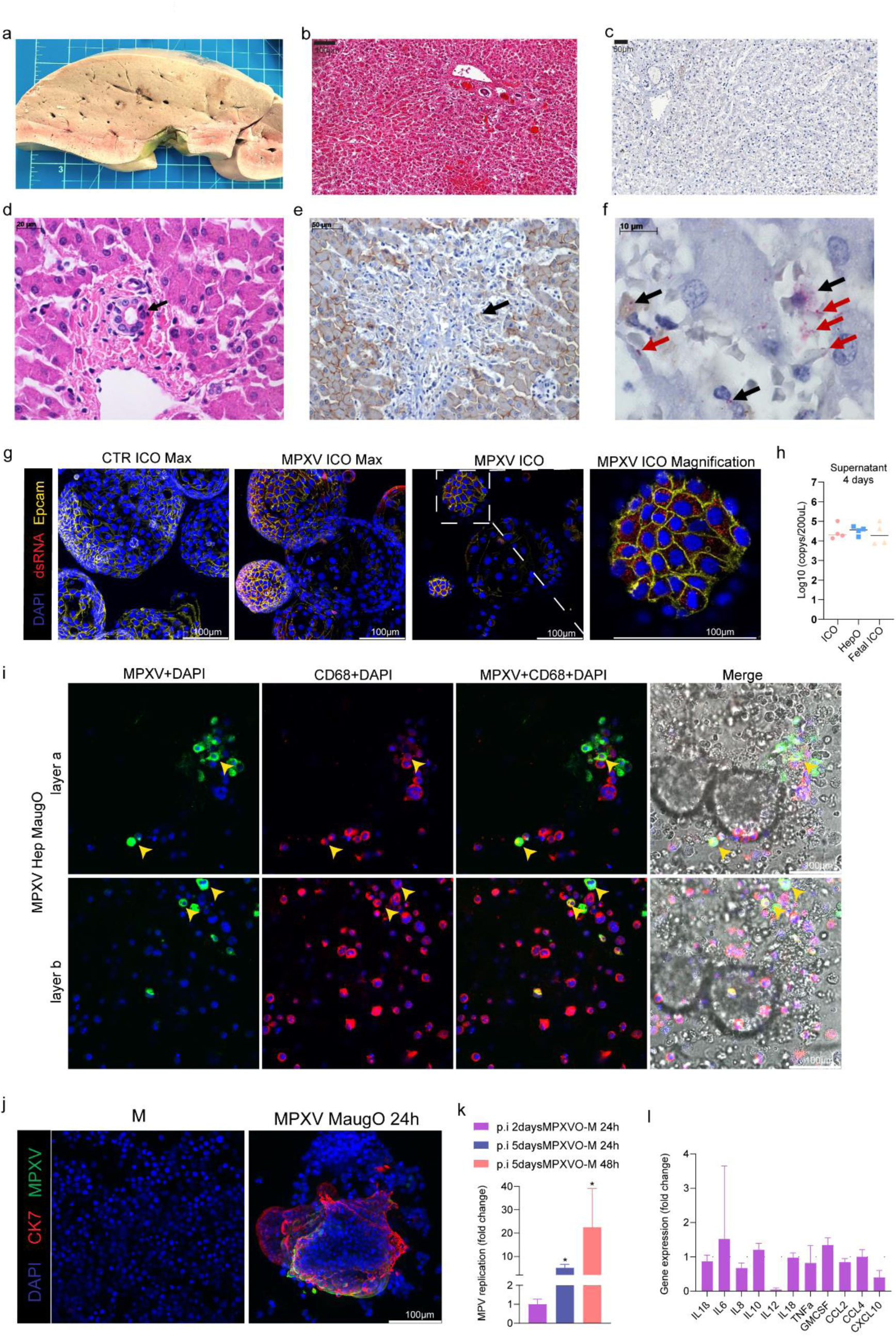
Characterizing MPXV infection in the liver of a patient and in organoids based models. a, Image of the liver from a patient died from disseminated mpox. Here we specifically focused on liver involvement in one case who developed MPXV-hepatitis. In the liver, we observed steatosis, diffuse congestion, focal subcapsular hemorrhages and scattered lobular nodules. b, Histology (H&E) of the liver section from a patient without MPXV infection (as a negative control). c, Immunohistochemical staining against the MPXV antigens showed negative signal in this negative control liver tissue, indicating the specificity of the antibody. d, Biliary duct exhibiting cholangiocytes with pykontic nuclei and hypereosinophiliccytoplasm (H&E). e, Focal MPXV-hepatitis (arrow), and the adjacent hepatocytes showing reduced expression of E-cadherin by immunohistochemical staining. f, Immunohistochemical staining of bacterial lipopolysaccharide (LPS) detected bacterial fragments in the cytoplasm of Kupffer cells (black arrows) or entire bacilli in the sinusoids (red arrows). g, Confocal imaging of MPXV infection in ICOs complementary to Fig. 7e. EpCAM (yellow): epithelial marker; dsRNA (red): MPXV double-stranded RNA; DAPI (blue): nuclear staining. h, Quantification of MPXV genomic DNA in the supernatant harvested from infected adult ICOs (O), hepatic differentiated ICOs (HepO), and fetal ICOs (FetalO) 4 days post-inoculation (n = 4), complementary to Fig. 7j. i, Confocal imaging shows MPXV infection in macrophages of MaugOs constructed from hepatic differentiated organoids, complementary to Fig. 8b. CD68 (red): macrophage marker; MPXV (green): staining against the virions; DAPI (blue): nuclear staining. j, Confocal imaging of uninfected macrophages (M) and ICOs infected with MPXV for 2days, and then integrated with macrophages for 24 hours (MPXV MaugO). CK7 (red): marker of biliary epithelium; MPXV (green): staining against the virions; DAPI (blue): nuclear staining. k, Quantification of MPXV viral RNA in MaugOs constructed by different approaches (n = 4). ICOs were infected with MPXV for 2 or 5 days, and then integrated with macrophages for 24 or 48 hours. Relating to Fig. 8h, ICOs were infected with MPXV for 5 days, and then integrated with macrophages for 24 or 48 hours. l, The expression of inflammatory genes in ICOs infected with MPXV for 2 days, and then integrated with macrophages for 24 hours (n = 4). (h, k, i) Data are mean ± SD. *p < 0.05; **p < 0.01; Mann-Whitney U test.

## Notes

### Competing Interest Statement

The authors have declared no competing interest.

### Author Declarations

Medical Ethical Council of the Erasmus MC

## References

1 Chiang, J. Y. L. & Ferrell, J. M. Bile Acid Metabolism in Liver Pathobiology. Gene Expr 18, 71–87 (2018). https://doi.org/GE379 [pii]10.3727/105221618X15156018385515

2 Ma, Z., de Man, R. A., Kamar, N. & Pan, Q. Chronic hepatitis E: Advancing research and patient care. J Hepatol 77, 1109–1123 (2022). https://doi.org/S0168-8278(22)00319-1 [pii]10.1016/j.jhep.2022.05.006

3 Zhao, Y. et al. Mechanistic insight of SARS-CoV-2 infection using human hepatobiliary organoids. Gut 72, 216–218 (2023). https://doi.org/gutjnl-2021-326617 [pii]10.1136/gutjnl-2021-326617

4 Luxenburger, H. & Thimme, R. SARS-CoV-2 and the liver: clinical and immunological features in chronic liver disease. Gut 72, 1783–1794 (2023). https://doi.org/gutjnl-2023-329623 [pii]10.1136/gutjnl-2023-329623

5 Wang, Y. et al. SARS-CoV-2 infection of the liver directly contributes to hepatic impairment in patients with COVID-19. J Hepatol 73, 807–816 (2020). https://doi.org/S0168-8278(20)30294-4 [pii]10.1016/j.jhep.2020.05.002

6 Li, X., Fan, C., Tang, J. & Zhang, N. Meta-analysis of liver injury in patients with COVID-19. Medicine (Baltimore) 102, e34320 (2023). https://doi.org/00005792-202307210-00038 [pii]10.1097/MD.0000000000034320

7 Muller, G. et al. Monkeypox virus in liver and spleen of child in Gabon. Lancet 1, 769–770 (1988). https://doi.org/S0140-6736(88)91580-2 [pii]10.1016/s0140-6736(88)91580-2

8 Uphoff, C. C., Pommerenke, C., Denkmann, S. A. & Drexler, H. G. Screening human cell lines for viral infections applying RNA-Seq data analysis. PLoS One 14, e0210404 (2019). https://doi.org/PONE-D-18-28319 [pii]10.1371/journal.pone.0210404

9 Han, Y., Yang, L., Lacko, L. A. & Chen, S. Human organoid models to study SARS-CoV-2 infection. Nat Methods 19, 418–428 (2022). https://doi.org/10.1038/s41592-022-01453-y [pii]10.1038/s41592-022-01453-y

10 Puschhof, J., Pleguezuelos-Manzano, C. & Clevers, H. Organoids and organs-on-chips: Insights into human gut-microbe interactions. Cell Host Microbe 29, 867–878 (2021). https://doi.org/S1931-3128(21)00150-5 [pii]10.1016/j.chom.2021.04.002

11 Casanova, J. L. & Abel, L. Mechanisms of viral inflammation and disease in humans. Science 374, 1080–1086 (2021). 10.1126/science.abj7965

12 Merad, M. & Martin, J. C. Pathological inflammation in patients with COVID-19: a key role for monocytes and macrophages. Nat Rev Immunol 20, 355–362 (2020). https://doi.org/10.1038/s41577-020-0331-4 [pii]331 [pii]10.1038/s41577-020-0331-4

13 Ginhoux, F. & Guilliams, M. Tissue-Resident Macrophage Ontogeny and Homeostasis. Immunity 44, 439–449 (2016). https://doi.org/S1074-7613(16)30063-2 [pii]10.1016/j.immuni.2016.02.024

14 Mosser, D. M. & Edwards, J. P. Exploring the full spectrum of macrophage activation. Nat Rev Immunol 8, 958–969 (2008). https://doi.org/nri2448 [pii]10.1038/nri2448

15 Li, Y. et al. Seasonal coronavirus infections trigger NLRP3 inflammasome activation in macrophages but is therapeutically targetable. Antiviral Res 216, 105674 (2023). https://doi.org/S0166-3542(23)00152-3 [pii]10.1016/j.antiviral.2023.105674

16 Paerewijck, O. & Lamkanfi, M. The human inflammasomes. Mol Aspects Med 88, 101100 (2022). https://doi.org/S0098-2997(22)00045-0 [pii]10.1016/j.mam.2022.101100

17 Coll, R. C., Schroder, K. & Pelegrin, P. NLRP3 and pyroptosis blockers for treating inflammatory diseases. Trends Pharmacol Sci 43, 653–668 (2022). https://doi.org/S0165-6147(22)00083-9 [pii]10.1016/j.tips.2022.04.003

18 Marsee, A. et al. Building consensus on definition and nomenclature of hepatic, pancreatic, and biliary organoids. Cell Stem Cell 28, 816–832 (2021). https://doi.org/S1934-5909(21)00162-4 [pii]10.1016/j.stem.2021.04.005

19 Cao, X., van den Hil, F. E., Mummery, C. L. & Orlova, V. V. Generation and Functional Characterization of Monocytes and Macrophages Derived from Human Induced Pluripotent Stem Cells. Curr Protoc Stem Cell Biol 52, e108 (2020). https://doi.org/CPSC108 [pii]10.1002/cpsc.108

20 Wang, W. et al. The RNA genome of hepatitis E virus robustly triggers an antiviral interferon response. Hepatology 67, 2096–2112 (2018). 10.1002/hep.29702

21 Beer, A. et al. Chronic Hepatitis E is associated with cholangitis. Liver Int 39, 1876–1883 (2019). https://doi.org/LIV14137 [pii]10.1111/liv.14137

22 Li, P. et al. Recapitulating hepatitis E virus-host interactions and facilitating antiviral drug discovery in human liver-derived organoids. Sci Adv 8, eabj5908 (2022). https://doi.org/abj5908 [pii]10.1126/sciadv.abj5908

23 Steiner, S. et al. SARS-CoV-2 biology and host interactions. Nat Rev Microbiol 22, 206–225 (2024). https://doi.org/10.1038/s41579-023-01003-z [pii]10.1038/s41579-023-01003-z

24 Guan, J., Fan, Y., Wang, S. & Zhou, F. Functions of MAP3Ks in antiviral immunity. Immunol Res 71, 814–832 (2023). https://doi.org/10.1007/s12026-023-09401-4 [pii]9401 [pii]10.1007/s12026-023-09401-4

25 Broutier, L. et al. Culture and establishment of self-renewing human and mouse adult liver and pancreas 3D organoids and their genetic manipulation. Nat Protoc 11, 1724–1743 (2016). https://doi.org/nprot.2016.097 [pii]10.1038/nprot.2016.097

26 Hamming, I. et al. Tissue distribution of ACE2 protein, the functional receptor for SARS coronavirus. A first step in understanding SARS pathogenesis. J Pathol 203, 631–637 (2004). https://doi.org/PATH1570 [pii]10.1002/path.1570

27 Shiratori, H. et al. THP-1 and human peripheral blood mononuclear cell-derived macrophages differ in their capacity to polarize in vitro. Mol Immunol 88, 58–68 (2017). https://doi.org/S0161-5890(17)30161-X [pii]10.1016/j.molimm.2017.05.027

28 Legrand, C. et al. Lactate dehydrogenase (LDH) activity of the cultured eukaryotic cells as marker of the number of dead cells in the medium [corrected]. J Biotechnol 25, 231–243 (1992). https://doi.org/0168-1656(92)90158-6 [pii]10.1016/0168-1656(92)90158-6

29 Chiang, J. Y. L. & Ferrell, J. M. Bile acid receptors FXR and TGR5 signaling in fatty liver diseases and therapy. Am J Physiol Gastrointest Liver Physiol 318, G554–G573 (2020). https://doi.org/GI-00223-2019 [pii]10.1152/ajpgi.00223.2019

30 Yin, X., Li, X. & Feng, Z. Role of Envelopment in the HEV Life Cycle. Viruses 8 (2016). https://doi.org/v8080229 [pii]viruses-08-00229 [pii]10.3390/v8080229

31 Hanafi, N. I., Mohamed, A. S., Sheikh Abdul Kadir, S. H. & Othman, M. H. D. Overview of Bile Acids Signaling and Perspective on the Signal of Ursodeoxycholic Acid, the Most Hydrophilic Bile Acid, in the Heart. Biomolecules 8 (2018). https://doi.org/biom8040159 [pii]biomolecules-08-00159 [pii]10.3390/biom8040159

32 Li, Y. et al. Hepatitis E virus infection activates NOD-like receptor family pyrin domain-containing 3 inflammasome antagonizing interferon response but therapeutically targetable. Hepatology 75, 196–212 (2022). https://doi.org/HEP32114 [pii]10.1002/hep.32114

33 Wang, Y. et al. Combating pan-coronavirus infection by indomethacin through simultaneously inhibiting viral replication and inflammatory response. iScience 26, 107631 (2023). https://doi.org/S2589-0042(23)01708-X [pii]107631 [pii]10.1016/j.isci.2023.107631

34 Group, R. C. et al. Dexamethasone in Hospitalized Patients with Covid-19. N Engl J Med 384, 693–704 (2021). https://doi.org/NJ202007173830003 [pii]10.1056/NEJMoa2021436

35 Li, P. et al. Clinical Features, Antiviral Treatment, and Patient Outcomes: A Systematic Review and Comparative Analysis of the Previous and the 2022 Mpox Outbreaks. J Infect Dis 228, 391–401 (2023). https://doi.org/7025706 [pii]jiad034 [pii]10.1093/infdis/jiad034

36 Mitja, O. et al. Mpox in people with advanced HIV infection: a global case series. Lancet 401, 939–949 (2023). https://doi.org/S0140-6736(23)00273-8 [pii]10.1016/S0140-6736(23)00273-8

37 Bruhn, P. J., Osterballe, L., Hillingso, J., Svendsen, L. B. & Helgstrand, F. Posttraumatic levels of liver enzymes can reduce the need for CT in children: a retrospective cohort study. Scand J Trauma Resusc Emerg Med 24, 104 (2016). https://doi.org/10.1186/s13049-016-0297-1 [pii]297 [pii]10.1186/s13049-016-0297-1

38 Duarte-Neto, A. N. et al. Main autopsy findings of visceral involvement by fatal mpox in patients with AIDS: necrotising nodular pneumonia, nodular ulcerative colitis, and diffuse vasculopathy. Lancet Infect Dis 23, 1218–1222 (2023). https://doi.org/S1473-3099(23)00574-1 [pii]10.1016/S1473-3099(23)00574-1

39 Li, P. et al. Mpox virus infection and drug treatment modelled in human skin organoids. Nat Microbiol 8, 2067–2079 (2023). https://doi.org/10.1038/s41564-023-01489-6 [pii]10.1038/s41564-023-01489-6

40 Li, P. et al. Mpox virus infects and injures human kidney organoids, but responding to antiviral treatment. Cell Discov 9, 34 (2023). https://doi.org/10.1038/s41421-023-00545-z [pii]545 [pii]10.1038/s41421-023-00545-z

41 Li, P. et al. Recapitulating infection, thermal sensitivity and antiviral treatment of seasonal coronaviruses in human airway organoids. EBioMedicine 81, 104132 (2022). https://doi.org/S2352-3964(22)00313-9 [pii]104132 [pii]10.1016/j.ebiom.2022.104132

42 Lamers, M. M. & Haagmans, B. L. SARS-CoV-2 pathogenesis. Nat Rev Microbiol 20, 270–284 (2022). https://doi.org/10.1038/s41579-022-00713-0 [pii]10.1038/s41579-022-00713-0

43 Vazquez-Armendariz, A. I. et al. Multilineage murine stem cells generate complex organoids to model distal lung development and disease. EMBO J 39, e103476 (2020). https://doi.org/EMBJ2019103476 [pii]10.15252/embj.2019103476

44 Vazquez-Armendariz, A. I. & Tata, P. R. Recent advances in lung organoid development and applications in disease modeling. J Clin Invest 133 (2023). https://doi.org/170500 [pii]10.1172/JCI170500

45 Recaldin, T. et al. Human organoids with an autologous tissue-resident immune compartment. Nature 633, 165–173 (2024). https://doi.org/10.1038/s41586-024-07791-5 [pii]7791 [pii]10.1038/s41586-024-07791-5

46 Huch, M. et al. Long-term culture of genome-stable bipotent stem cells from adult human liver. Cell 160, 299–312 (2015). https://doi.org/S0092-8674(14)01566-9 [pii]10.1016/j.cell.2014.11.050

47 Roos, F. J. M. et al. Human branching cholangiocyte organoids recapitulate functional bile duct formation. Cell Stem Cell 29, 776–794 e713 (2022). https://doi.org/S1934-5909(22)00164-3 [pii]10.1016/j.stem.2022.04.011

48 Castejon-Ramirez, S., Pennington, J., Beene, H., Hysmith, N. & Ost, S. A Case of Neonatal Monkeypox Treated With Oral Tecovirimat. Pediatrics (2023). https://doi.org/196275 [pii]10.1542/peds.2023-061198

49 Yakubovsky, M. et al. Mpox Presenting as Proctitis in Men Who Have Sex With Men. Clin Infect Dis 76, 528–530 (2023). https://doi.org/6692817 [pii]10.1093/cid/ciac737

50 Arfi, V. et al. Characterization of the early steps of infection of primary blood monocytes by human immunodeficiency virus type 1. J Virol 82, 6557–6565 (2008). https://doi.org/JVI.02321-07 [pii]2321-07 [pii]10.1128/JVI.02321-07

51 MacParland, S. A. et al. Single cell RNA sequencing of human liver reveals distinct intrahepatic macrophage populations. Nat Commun 9, 4383 (2018). https://doi.org/10.1038/s41467-018-06318-7 [pii]6318 [pii]10.1038/s41467-018-06318-7

52 Hakim, M. S. et al. The global burden of hepatitis E outbreaks: a systematic review. Liver Int 37, 19–31 (2017). 10.1111/liv.13237

53 Davis, H. E., McCorkell, L., Vogel, J. M. & Topol, E. J. Long COVID: major findings, mechanisms and recommendations. Nat Rev Microbiol 21, 133–146 (2023). https://doi.org/10.1038/s41579-022-00846-2 [pii]846 [pii]10.1038/s41579-022-00846-2

54 Wong, A. C. et al. Serotonin reduction in post-acute sequelae of viral infection. Cell 186, 4851–4867 e4820 (2023). https://doi.org/S0092-8674(23)01034-6 [pii]10.1016/j.cell.2023.09.013

55 Pan, Q. et al. Mobilization of hepatic mesenchymal stem cells from human liver grafts. Liver Transpl 17, 596–609 (2011). 10.1002/lt.22260

56 Ettayebi, K. et al. Replication of human noroviruses in stem cell-derived human enteroids. Science 353, 1387–1393 (2016). https://doi.org/science.aaf5211 [pii]10.1126/science.aaf5211

57 Reese, V. C., Oropeza, C. E. & McLachlan, A. Independent activation of hepatitis B virus biosynthesis by retinoids, peroxisome proliferators, and bile acids. J Virol 87, 991–997 (2013). https://doi.org/JVI.01562-12 [pii]01562-12 [pii]10.1128/JVI.01562-12

58 Chhatwal, P. et al. Bile acids specifically increase hepatitis C virus RNA-replication. PLoS One 7, e36029 (2012). https://doi.org/PONE-D-11-19324 [pii]10.1371/journal.pone.0036029

59 Lloyd, G., Atkinson, T. & Sutton, P. M. Effect of bile salts and of fusidic acid on HIV-1 infection of cultured cells. Lancet 1, 1418–1421 (1988). https://doi.org/S0140-6736(88)92236-2 [pii]10.1016/s0140-6736(88)92236-2

60 Winkler, E. S. et al. The Intestinal Microbiome Restricts Alphavirus Infection and Dissemination through a Bile Acid-Type I IFN Signaling Axis. Cell 182, 901–918 e918 (2020). https://doi.org/S0092-8674(20)30806-0 [pii]10.1016/j.cell.2020.06.029

61 Luo, L. et al. Chenodeoxycholic Acid from Bile Inhibits Influenza A Virus Replication via Blocking Nuclear Export of Viral Ribonucleoprotein Complexes. Molecules 23 (2018). https://doi.org/molecules23123315 [pii]molecules-23-03315 [pii]10.3390/molecules23123315

62 Kong, F., Saif, L. J. & Wang, Q. Roles of bile acids in enteric virus replication. Anim Dis 1, 2 (2021). https://doi.org/3 [pii]10.1186/s44149-021-00003-x

63 Haselow, K. et al. Bile acids PKA-dependently induce a switch of the IL-10/IL-12 ratio and reduce proinflammatory capability of human macrophages. J Leukoc Biol 94, 1253–1264 (2013). https://doi.org/jlb.0812396 [pii]10.1189/jlb.0812396

64 Arbel, R. et al. Nirmatrelvir Use and Severe Covid-19 Outcomes during the Omicron Surge. N Engl J Med 387, 790–798 (2022). https://doi.org/NJ202208243870901 [pii]10.1056/NEJMoa2204919

65 Desai, A. N. et al. Compassionate Use of Tecovirimat for the Treatment of Monkeypox Infection. JAMA 328, 1348–1350 (2022). https://doi.org/2795743 [pii]jld220061 [pii]10.1001/jama.2022.15336

66 Arias, C. A., Miller, W. R., Olsen, R., Gollihar, J. & Armstrong, A. The response of mpox-associated inflammatory syndrome to steroid therapy. Lancet Infect Dis 23, e323–e324 (2023). https://doi.org/S1473-3099(22)00876-3 [pii]10.1016/S1473-3099(22)00876-3

67 de Almeida, L. et al. Identification of immunomodulatory drugs that inhibit multiple inflammasomes and impair SARS-CoV-2 infection. Sci Adv 8, eabo5400 (2022). https://doi.org/abo5400 [pii]10.1126/sciadv.abo5400

68 Malireddi, R. K. S. et al. Inflammatory cell death, PANoptosis, screen identifies host factors in coronavirus innate immune response as therapeutic targets. Commun Biol 6, 1071 (2023). https://doi.org/10.1038/s42003-023-05414-9 [pii]5414 [pii]10.1038/s42003-023-05414-9

69 Gautam, A. et al. Necroptosis blockade prevents lung injury in severe influenza. Nature 628, 835–843 (2024). https://doi.org/10.1038/s41586-024-07265-8 [pii]10.1038/s41586-024-07265-8

