## Supplemental information for "Macrophage-augmented organoids recapitulate the complex pathophysiology of viral diseases and enable development of multitarget therapeutics"

### Supplementary file

#### Table S1: Key resources used in this study

| REAGENT or RESOURCE | SOURCE | IDENTIFIER |
|---|---|---|
| **Antibodies against** | | |
| IL-1β (D3U3E), rabbit | Cell Signaling Technology | 12703 |
| Cleaved IL-1β, rabbit | Cell Signaling Technology | 83186s |
| NLRP3, rabbit | ThermoFisher Scientific | PA5-20838 |
| dsRNA, mouse | SCICONS | 10010200 |
| HEV, MS x 1E6 | Sigma-Aldrich Chemie BV | MAB8002 |
| ORF2-specific rabbit hyperimmune serum | Group R. Ullrich, Friedrich Loeffler Institute, Germany | N/A |
| β-actin, mouse | Santa Cruz Biotechnology | sc-47778 |
| IRDye® 680RD Goat anti-Mouse IgG (H + L) | Westburg BV | 926-68070 |
| IRDye® 800CW Goat anti-Rabbit IgG (H + L) | Westburg BV | 926-32211 |
| Epcam, rabbit | Abcam | ab71916 |
| CD68, mouse | ThermoFisher Scientific | 14-0688-82 |
| Cleaved caspase 3, rabbit | Cell Signaling Technology | 9664S |
| SARS-CoV-2-nucleocapsid protein, rabbit | Thermo Fisher Scientific | MA5-29981 |
| Albumin, rabbit | Thermo Fisher Scientific | MA5-29022 |
| Vaccinia virus Lister Strain (FITC), rabbit | Abbexa | abx023199 |
| CK7 (KRT7), mouse | DAKO | AKO: M7018 |
| Vaccinia virus, Rabbit | Thermo Fisher Scientific | PA1-7258 |

| | | |
|---|---|---|
| CD68 | DAKO | M0876 |
| E-cadherin | Cell Marque | EP700Y |
| LPS | Abcam | Ab35654 |
| Alexa Fluor 488 Goat – anti-rabbit | Thermo Fisher Scientific | A32731 |
| Alexa Fluor 555 Goat anti-Rabbit | ThermoFisher Scientific | A-21428 |
| Alexa Fluor 594 Goat anti-Mouse | ThermoFisher Scientific | A32742 |
| Alexa Fluor 647 Goat anti-Rabbit | Thermo Fisher Scientific | A-21245 |
| **Virus strains** | | |
| HEV, Kernow-C1 p6 clone, JQ679013 | https://www.ncbi.nlm.nih.gov/nuccore/JQ679013 | N/A |
| SARS-CoV-2 Beta Netherlands | Erasmus Medical Center | GenBank: MT270101 |
| Monkeypox virus | European Virus Archive | Ref-SKU: 010V-04721 |
| **Chemicals** | | |
| Matrigel | Corning | 356231 |
| Immobilon® Block - FL (Fluorescent Blocker) | Merck Chemicals BV | WBAVDFL01 |
| Intercept® (PBS) Blocking Buffer | LI-COR | 927-70010 |
| Immobilon-FL PVDF, 0.45 µm, 8.5 cm x 10 m roll | Merck Chemicals BV | IPFL85R |
| PrimeScript™ RT Master Mix (Perfect Real Time) | Takara Bio Europe S.A.S. | RR036A |
| Methanol | Boom | 72,032,213.2500 |
| TrypLE | ThermoFisher Scientific | 12,604,013 |
| HEPES | Lonza/Fisher Scientific | BE17-737E |
| N-2 supplement (50x) | Gibco/ThermoFisher Scientific | 15,410,294 |

| | | |
|---|---|---|
| B27 supplement 50x without vitamin A | Gibco/ThermoFisher Scientific | 12,587,001 |
| Gastrin I | Sigma-Aldrich | G9145 |
| FGF10 | Peprotech | 100–26 |
| HGF | Tebu-bio | 167,100-39-0500 |
| EGF | Peprotech | AF-100-15 |
| A83-01 | Cayman Chemical/Sanbio | 9,001,799–25 |
| Nicotinamide | Sigma-Aldrich | N0636-100G |
| Forskolin | Sigma-Aldrich | F6886 |
| RSPO1 | Home made | N/A |
| Y-27632 | MedChem Express/Bioconnect | HY-10583_10mg |
| DAPT Selleck | Chemicals/Bioconnect | S2215_10mg |
| Human BMP-7 | Tebu-bio | 167,120-03P-B |
| Dexamethasone | Sigma-Aldrich | D4902-100MG |
| DMEM high glucose w/Na pyruvate w/ Stable glutamine | VWR International BV | L0193-500 |
| PMA(12-O-Tetradecanoylphorbol 13-acetate) | Sigma-Aldrich Chemie BV | P1585 |
| RPMI 1640 (STABLE GLUTAMINE) | Westburg BV | L0498-500 |
| **Organoids, primary cells and cell lines** | | |
| ICOs (intrahepatic cholangiocyte organoids) (n = 11; from adult donors) | Erasmus University Medical Center | N/A |
| Primary macrophages (pooled from different donors) | Sanquin | N/A |
| Hepatic macrophages (n = 9) | Erasmus University Medical Center | N/A |

| | | |
|---|---|---|
| HepO (hepatocyte-differentiated ICOs) (n = 2) | Erasmus University Medical Center | N/A |
| FLO (Fetal ICOs) (n = 1) | Erasmus University Medical Center | N/A |
| Human induced pluripotent stem cell line | The Allen Cell Collection | AICS-0061-036 |
| THP-1 | Erasmus University Medical Center | N/A |
| Huh7 | Erasmus University Medical Center | N/A |
| Vero-E6 | Erasmus University Medical Center | N/A |
| **Software** | | |
| Adobe Illustrator CC | Adobe | https://www.adobe.com/products/illustrator.html |
| GraphPad Prism 8.0 | GraphPad | Graphpad Software |
| ImageJ | NiH | https://ImageJ.nih.gov/ij/ |
| Rstudio | Rstudio | https://www.rstudio.com/products/rstudio/download/ |
| FlowJo | BD | https://www.flowjo.com/solutions/flowjo/downloads |
| BD FACSDiva Software v9.1 | BD | https://www.bdbiosciences.com/en-us/products/software/instrument-software/bd-facsdiva-software |

**Table S2: Donor information for generating organoids and culturing hepatic macrophages**

| **Liver tissues for generating Organoids** | |
|---|---|
| Donor | Gender |
| 1 | Male |
| 2 | Male |
| 3 | Male |
| 4 | Female |
| 5 | Female |
| 6 | Female |
| 7 | Female |
| 8 | Female |
| 9 | Female |
| 10 | Male |
| 11 | Female |
| **Hepatic macrophages from perfusates** | |
| Donor | Gender |
| 1 | Male |
| 2 | Female |
| 3 | Female |
| 4 | male |
| 5 | Female |
| 6 | Female |
| 7 | Male |
| 8 | Female |
| 9 | Female |

**Table S3: Primer sequences used in this study**

| Gene | Sequence (5’-3’) |
|---|---|
| HEV sense | GGTGGTTTCTGGGGTGAC |
| HEV anti sense | AGGGGTTGGTTGGATGAA |
| GAPDH sense | GTCTCCTCTGACTTCAACAGCG |
| GAPDH anti sense | ACCACCCTGTTGCTGTAGCCAA |
| IL-1β sense | CCACAGACCTTCCAGGAGAATG |
| IL-1β anti sense | GTGCAGTTCAGTGATCGTACAGG |
| IL-6 sense | AGACAGCCACTCACCTCTTCAG |
| IL-6 anti sense | TTCTGCCAGTGCCTCTTTGCTG |
| IL-8 sense | GAGAGTGATTGAGAGTGGACCAC |
| IL-8 anti sense | CACAACCCTCTGCACCCAGTTT |
| IL-10 sense | TCTCCGAGATGCCTTCAGCAGA |
| IL-10 anti sense | TCAGACAAGGCTTGGCAACCCA |
| IL-12 sense | GACATTCTGCGTTCAGGTCCAG |
| IL-12 anti sense | CATTTTTGCGGCAGATGACCGTG |
| IL-18 sense | GATAGCCAGCCTAGAGGTATGG |
| IL-18 anti sense | CCTTGATGTTATCAGGAGGATTCA |
| TNF α sense | CTCTTCTGCCTGCTGCACTTTG |
| TNF α anti sense | ATGGGCTACAGGCTTGTCACTC |
| GM-CSF sense | GGAGCATGTGAATGCCATCCAG |
| GM-CSF anti sense | CTGGAGGTCAAACATTTCTGAGAT |
| CCL-2 sense | AGAATCACCAGCAGCAAGTGTCC |
| CCL-2 anti sense | TCCTGAACCCACTTCTGCTTGG |
| CCL-4 sense | GCTTCCTCGCAACTTTGTGGTAG |

| | |
|---|---|
| CCL-4 anti sense | GGTCATACACGTACTCCTGGAC |
| CXCL-10 sense | GGTGAGAAGAGATGTCTGAATCC |
| CXCL-10 anti sense | GTCCATCCTTGGAAGCACTGCA |
| iNOS sense | GCTCTACACCTCCAATGTGACC |
| iNOS anti sense | CTGCCGAGATTTGAGCCTCATG |
| Arg-1 sense | TCATCTGGGTGGATGCTCACAC |
| Arg-1 anti sense | GAGAATCCTGGCACATCGGGAA |
| SARS-CoV-2 sense | CAATGGTTTAACAGGCACAGG |
| SARS-CoV-2 anti sense | CTCAAGTGTCTGTGGATCACG |
| MPXV sense | GGCTCTTCTATCAACCACA |
| MPXV anti sense | AGTCATTATCTCCTCCTCCA |
| Albumin sense | GATGAGATGCCTGCTGACTTGC |
| Albumin anti sense | CACGACAGAGTAATCAGGATGCC |
| LGR5 sense | CCTGCTTGACTTTGAGGAAGACC |
| LGR5 anti sense | CCAGCCATCAAGCAGGTGTTCA |
| G6PC sense | GCTGTGATTGGAGACTGGCTCA |
| G6PC anti sense | GTCCAGTCTCACAGGTTACAGG |

**Supplementary Video 1:** Visualization the formation of macrophage-augmented organoids. THP-1 macrophages were pre-labeled with CFSE dye (green), and the organoids were mechanically dissociated into small fragments.

**Supplementary Video 2:** 3D reconstruction of MaugOs, commentary to Fig. 1f. DAPI (blue), nuclear staining; Epcam marked organoids (red); CFSE labeled macrophages (green).

**Supplementary Video 3:** 3D reconstruction of MaugOs, commentary to Fig. 1g. Epcam marked organoids (red); CFSE labeled macrophages (green).
